# Improved estimates of COVID-19 correlates of protection, antibody decay and vaccine efficacy waning: a joint modelling approach

**DOI:** 10.1101/2024.07.02.24309776

**Authors:** Daniel J. Phillips, Maria D. Christodoulou, Shuo Feng, Andrew J. Pollard, Merryn Voysey, David Steinsaltz

**Affiliations:** Department of Statistics, University of Oxford, Oxford, UK; Oxford Vaccine Group, Department of Paediatrics, University of Oxford, Oxford, UK; NIHR Oxford Biomedical Research Centre, Oxford, UK July 2, 2024

**Author notes:** Correspondence to Daniel J. Phillips,.

## Abstract

Reliable estimation of the relationship between COVID-19 antibody levels at the time of exposure and the risk of infection is crucial to inform policy decisions on vaccination regimes. We fit a joint model of anti-spike IgG antibody decay and risk of COVID-19 infection to data from a randomized efficacy trial of the ChAdOx1 nCoV-19 vaccine. Our model improves upon previous analyses by accounting for measurement error, decay in antibody levels and variation between different individuals. We estimated correlates of protection, antibody decay, and vaccine efficacy waning. Increased anti-spike IgG antibody levels at the time of exposure correlate with increased vaccine-induced protection. We estimated vaccine efficacy against symptomatic COVID-19 infection of 88.1% (95% CrI: 77.2, 93.6) at day 35, waning to 60.4% (44.6, 71.0) at day 189 since the second dose. We report that longer intervals between the first and second vaccine dose give lasting increased protection, and observe lower efficacy in individuals aged ≥70 years from around 3 months after second dose. Our methods can be used in future vaccine trials to help inform the timings and priority of vaccine administration against novel diseases.

## Introduction

Correlates of protection, i.e. immune markers which relate to the risk of a disease outcome, are crucial to understanding the biological mechanisms for protection against infection after a given treatment. Most studies consider how immune responses at a fixed time soon after treatment correlate with risk of a disease outcome. However, it is the immune markers present at the time of exposure which protect against infection. Modelling immune levels over time and relating these to vaccine-induced protection, allows a clearer understanding of the relationship between immune responses and protection, and avoids potential biases due to waning of immune levels between the measurement and exposure. This further provides a natural framework to investigate the effect of factors such as age, comorbidities and dose schedules on immune response and protection, and to estimate changes in levels of protection over time.

In response to the COVID-19 pandemic, the (Oxford-AstraZeneca) ChAdOx1 nCoV-19 (AZD1222) vaccine has been widely administered in the UK and worldwide, alongside other COVID-19 vaccines. Binding and neutralising antibodies have been shown to be correlates of protection for ChAdOx1 nCoV-19 [1, 2] and other COVID-19 vaccines [3, 4, 5, 6, 7]. These studies all considered how initial peak antibody responses correlate with the risk of future COVID-19 infection, as this is the timepoint most relevant for vaccine licensure. If used to understand how antibody levels at the time of exposure relate to risk of infection, the above approaches may be biased, as they do not account for the waning of antibody levels between the time of the antibody measurement and exposure [8]. Other studies focussed on the relationship between antibody levels at exposure and the risk of subsequent infection [9, 10, 11]. Wei et al. (2022, 2023) used observational data to estimate how risk of infection after vaccination relates to a recent antibody measurement [9, 10]. However the use of observational data may cause their results to be biased. Follmann et al. (2023) modelled antibody decay over time after vaccination, relating the risk of infection to the predicted antibody level at the time of exposure [11]. However, Follmann et al. assume the same rate of decay for all individuals, which may decrease power and lead to bias. Further, all the above studies on correlates of protection use only one antibody measurement per individual. This means they are unable to account for measurement error, which will likely introduce bias to the results [12].

Vaccine efficacy (VE) has been shown to wane over time after two doses of the ChAdOx1 nCoV-19, BNT162b2, and mRNA-1273 vaccines in the UK and elsewhere, against the Alpha variant [13] and the Delta variant [13, 14, 15, 16, 17, 18]; as well as after a third booster dose [18, 19]. These studies all use observational data, in test-negative or cohort designs, which may introduce bias due to unmeasured confounders affecting both the probability of vaccination and of the clinical outcome (e.g. infection) [20, 21]. Randomised controlled trials are free of such bias, however sample sizes tend to be much smaller, restricting power for analysis of efficacy waning. Including antibody data in a model for vaccine efficacy waning may increase the power of the analysis [22, 8].

Joint models of longitudinal and time-to-event data are a powerful tool to investigate how longitudinal biomarker trajectories relate to the risk of health- or disease-related events [8]. The method has been applied to data on various diseases, especially disease progression in HIV/AIDS [23] and cancer trials [8]. See Ibrahim, Chu and Chen (2010) [8] for an introductory review, and Gould et al. (2015) [24] for a review with more detail on implementation methods and packages. Joint modelling has been shown to reduce bias and increase power in estimating the relationship between a longitudinal biomarker and clinical event [8, 25], when compared with methods which use observed biomarkers directly to predict risk. This is because the approach accounts for biomarker measurement error, as well as changes in the biomarker over time. Joint models have been applied to understand how viral load [26] and changes in biomarkers [27] relate to risk of mortality in COVID-19 patients, and to predict their risk of mortality from biomarker data [28]. They have also previously been used to understand how vaccine-induced immune responses relate to risk of AIDS/death in HIV patients [29], and risk of relapse/death in cancer patients [30]. We are not aware of any previous work applying joint modelling to understand how vaccine-induced responses relate to risk of infection with COVID-19 or any other infectious disease.

We implement a two-stage joint model for COVID-19 antibody decay and risk of COVID-19 infection after vaccination using multiple imputation [31]. Our model accounts for antibody decay over time, individual differences in decay rates and antibody measurement error. We apply our model to data from a randomised vaccine efficacy trial of two doses of the ChAdOx1 nCoV-19 vaccine [32, 33] to investigate three main outcomes (COV002). Firstly, we model the waning of anti-spike IgG antibody levels over time. Secondly, we estimate correlates of protection - the relationship between anti-spike IgG antibody levels at the time of exposure and the risk of COVID-19 infection. Thirdly, we estimate waning vaccine efficacy over time, and investigate covariate effects on antibody and vaccine efficacy waning.

## Results

We used data from the COV002 trial [32, 33] to assess antibody waning, vaccine efficacy (VE) waning, and correlates of protection against any COVID-19 infection and symptomatic COVID-19 infection at the time of exposure. COV002 is a randomized single-blind vaccine efficacy trial of the ChAdOx1 nCoV-19 vaccine, conducted in the United Kingdom. Participants were considered at risk of infection from 21 days after their second dose of vaccine, over a period from 18 July 2020 to 30 June 2021. This analysis includes 4605 ChAdOx1 nCoV-19 vaccinated individuals (vaccine arm) and 4423 individuals who received a MenACWY control vaccine (control arm). Supplementary Table 1 summarises baseline characteristics for the vaccine and control arm. Supplementary Fig. 1 summarises exclusions for the two groups. In the vaccine and control arm, 207 (4.5%) and 377 (8.5%) tested positive for COVID-19 respectively, and 71 (1.5%) and 217 (4.9%) were primary symptomatic COVID-19 cases respectively (Supplementary Table 1).

### Waning anti-spike IgG antibody levels

Blood samples were taken at study visits in windows centred at 28 days, 90 days and 182 days after the second dose. We refer to these as the PB28 (post-boost + 28 days), PB90 and PB182 study visits respectively. Samples were sent for anti-spike IgG antibody testing by case-cohort sampling. Samples taken after a reported COVID-19 infection were excluded from analysis. Observed antibody responses differed between those who did not return a positive COVID-19 test during the at risk period (non-cases), all who later tested positive (cases) and those who tested positive with primary symptoms (symptomatic cases). At the PB28, PB90 and PB182 study visits, antibody observations were available from 1155 (26.3%), 519 (11.8%), 58 (1.3%) non-cases, 173 (83.6%), 64 (30.9%), 1 (0.5%) cases, and 59 (83.1%), 23 (32.4%), 1 (1.4%) symptomatic cases respectively (Supplementary Table 2). At the PB28, PB90 and PB182 study visits, observed antibody levels had a median (interquartile range (IQR)) of 219 (119, 383), 115 (66, 206), 65 (45, 117) BAU/mL among non-cases, 197 (103, 322), 93 (47, 168), 30 (NA – only one measurement) BAU/mL among cases and 165 (100, 276), 78 (62, 178), 30 (NA – only one measurement) BAU/mL among symptomatic cases respectively. Fig. 1a shows the observed antibody levels against time, plotted on a log scale. The figure indicates (i) log antibody levels decay linearly, (ii) different individuals have different levels of initial log antibody response and (iii) may have different rates of decay.

**Figure 1:**
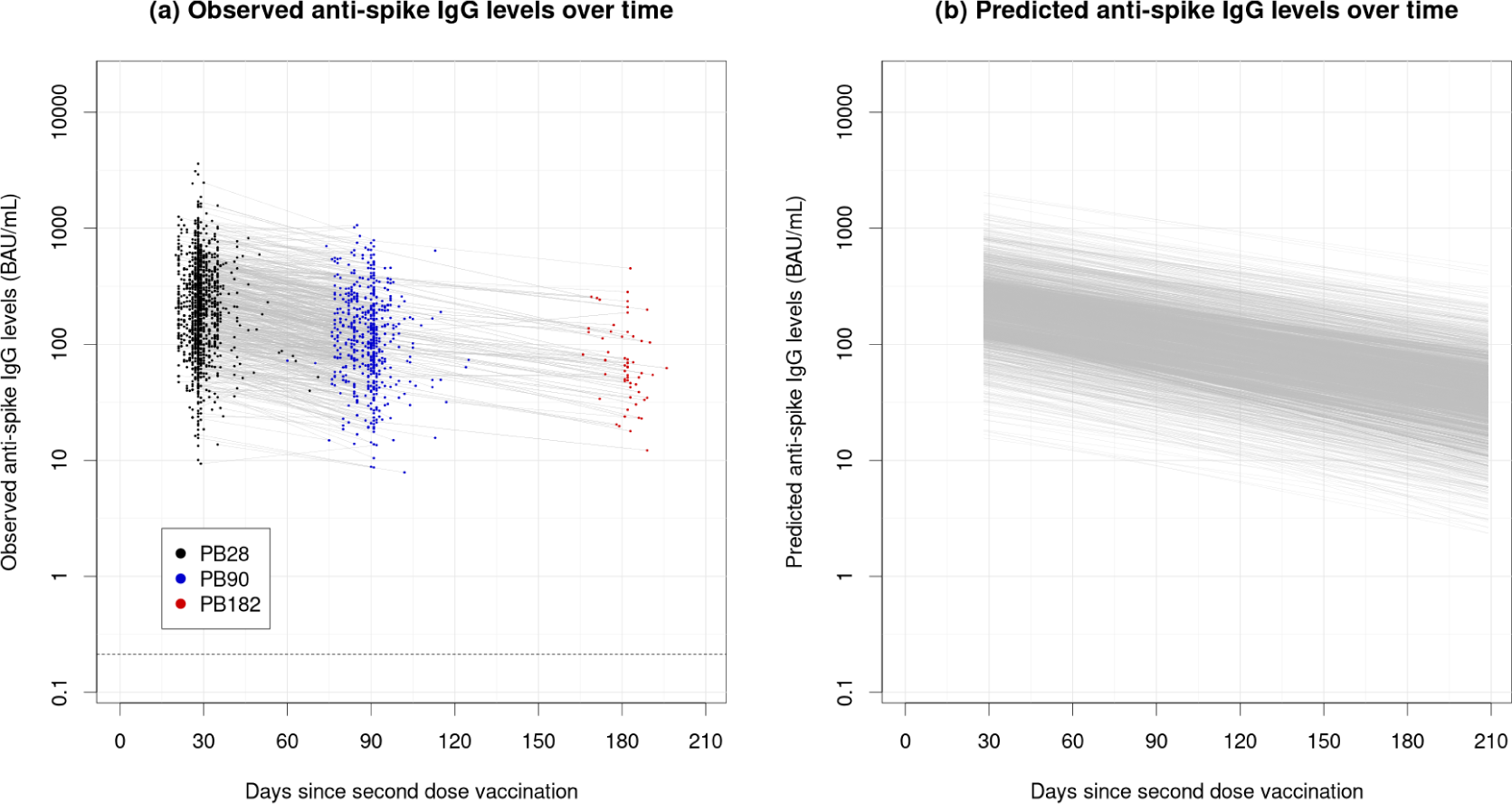
Observed and estimated anti-spike IgG antibody levels over time since vaccination. **(a)** Observed anti-spike IgG antibody levels over time since vaccination. Each point represents an observation, and consecutive observations from the same individual are connected by a line. Black, blue and red points represent observations taken at the PB28, PB90 and PB182 study visits respectively. The dashed horizontal line shows the lower limit of quantification (LLOQ), 0.21 BAU/mL. **(b)** The posterior mean estimate of the latent true antibody levels over time for each individual. Note 1b represent estimates of the latent true antibody levels which are free from measurement error, whereas the observed antibody levels in 1a are measured with error, so show higher variability.

In total, 923 antibody observations were available from 853 (18.5%) control participants. Among control participants, antibody levels had a median (IQR) of 0.3 (< 0.21 (lower limit of quantification (LLOQ)), 0.5) BAU/mL (Supplementary Fig. 2, Supplementary Table 2). In our subsequent modelling we did not model antibody levels in the control arm, nor include an antibody effect on risk for control participants.

We modelled the decay in antibody levels over time among the vaccinated participants, allowing different initial antibody responses and rates of decay for different individuals (Methods). Predicted median anti-spike IgG levels at day 28, 90 and 182 were 217 (95% credible interval (CrI): 208, 227), 119 (113, 126), 49 (45, 54) BAU/mL respectively. The median half-life of the anti-spike IgG antibody levels was 74 days (95% CrI: 69, 79). Fig. 1b shows the predicted antibody levels over time for each individual in the study on a log-scale.

Predicted antibody levels and rates of decay varied among individuals. The estimated 25% and 75% quartiles for the antibody response at day 28 were given by 124 (95% CrI: 117, 130) and 378 (359, 398) respectively (Supplementary Fig. 9). The half-life of the antibody levels among individuals in the study had an estimated 25% and 75% quartile of 62 days (95% CrI: 58, 67) and 87 days (79, 96). Among non-cases, the median (IQR) predicted anti-spike IgG level at day 28 was 219 (95% CrI: 209, 229), among cases 192 (175, 209) and among symptomatic cases 169 (146, 196). The median half-life was 74 (95% CrI: 70, 79) among non-cases, 60 (52, 69) among cases and 54 (45, 69) among symptomatic cases.

### Correlates of protection

We modelled how the protective effect of the vaccine relates to the modelled antibody levels over time. The instantaneous risk of COVID-19 infection for a given individual in our model depends on: (i) the current background levels of COVID-19, (ii) covariates which may affect risk of COVID-19 infection (iii) the modelled level of antibody 7 days prior, and (iv) a non-antibody effect due to vaccination (Methods). We assume a 7 day gap between initial exposure to COVID-19 and a positive test being reported.

Supplementary Fig. 3-6 show an estimate of the baseline hazard of infection during the trial period, the number of participants at risk over time since vaccination, the timings of antibody samples, and the timings of COVID-19 infections in the study respectively. Differences in baseline risk of infection due to different covariates are reported in Supplementary Fig. 7 and Supplementary Table 6.

We assessed how predicted antibody level at the time of exposure correlates with protection against COVID-19 infection. Predicted anti-spike IgG titer was correlated with protection. Fig. 2 shows the relationship between VE and anti-spike IgG antibody level predicted by our model. Anti-spike IgG values of 10, 100 and 1000 BAU/mL represent the 0.0%, 17.7% and 97.1% quantiles of predicted antibody levels at day 28 post second dose, and the 5.4%, 78.1% and 100% quantiles of predicted levels at day 182 respectively. At anti-spike IgG levels of 10, 100 and 1000 BAU/mL, the VE against symptomatic infection was 36.1% (95% CrI: −20.7, 63.0), 76.1% (64.4, 88.7), 100% (97.3, 100) respectively. We observe lower efficacy against any COVID-19 infection at the same antibody levels, 14.3% (95% CrI: −22.5, 38.1), 55.3% (44.4, 66.3), 99.9% (96.2, 100) respectively. Vaccine efficacy of 50%, 70%, and 90% against symptomatic infection was achieved at anti-spike IgG levels of 32 (95% CrI: 0, 57), 78 (51, 139), 182 (105, 507) BAU/mL respectively (Supplementary Table 7). Against any COVID-19 infection higher antibody levels were required for the same efficacy, 84 (95% CrI: 59, 125), 156 (109, 285), 311 (194, 662) BAU/mL respectively.

**Figure 2:**
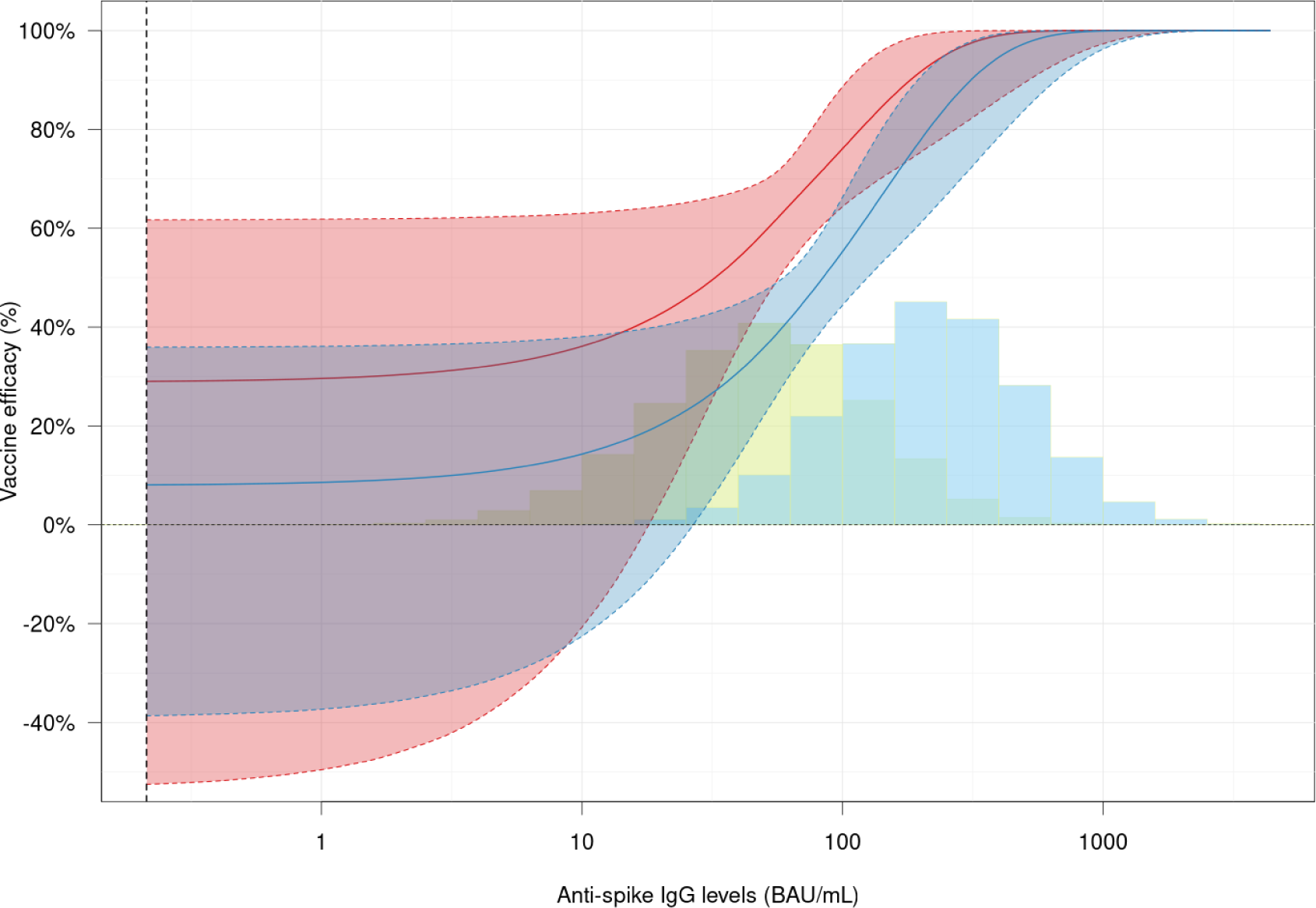
Vaccine efficacy as a function of anti-spike IgG levels. VE (%) plotted against anti-spike IgG antibody levels (BAU/mL) on a log scale. The red and blue curves and shaded regions show the posterior median and associated 95% credible intervals for VE against primary symptomatic COVID-19 infection, and against all NAAT+ positive tests respectively. The vertical dotted line shows the lower limit of quantification (LLOQ), 0.21 BAU/mL. The blue histogram shows the distribution of the predicted anti-spike IgG antibody levels in the study at 28 days since the second dose, and the green histogram at 182 days post second dose. The two histograms overlap. The heights of the histograms are not to scale.

### Waning vaccine efficacy

As the antibody levels decayed over time, the vaccine efficacy waned. Fig. 3 shows the mean VE over time since the second dose, averaged over all individuals in the study. We report efficacy 7 days after the planned study visits. The efficacy at day 35 after the second dose was estimated to be 88.1% (95% CrI: 77.2, 93.6) against symptomatic infection and 76.0% (64.3, 83.7) against all COVID-19 infection. The efficacy waned to 77.7% (68.5, 84.1) and 60.8% (51.4, 68.4) at day 97 and 60.4% (44.6, 71.0) and 39.6% (26.1, 50.2) at day 189 against symptomatic infection and all COVID-19 infection respectively.

**Figure 3:**
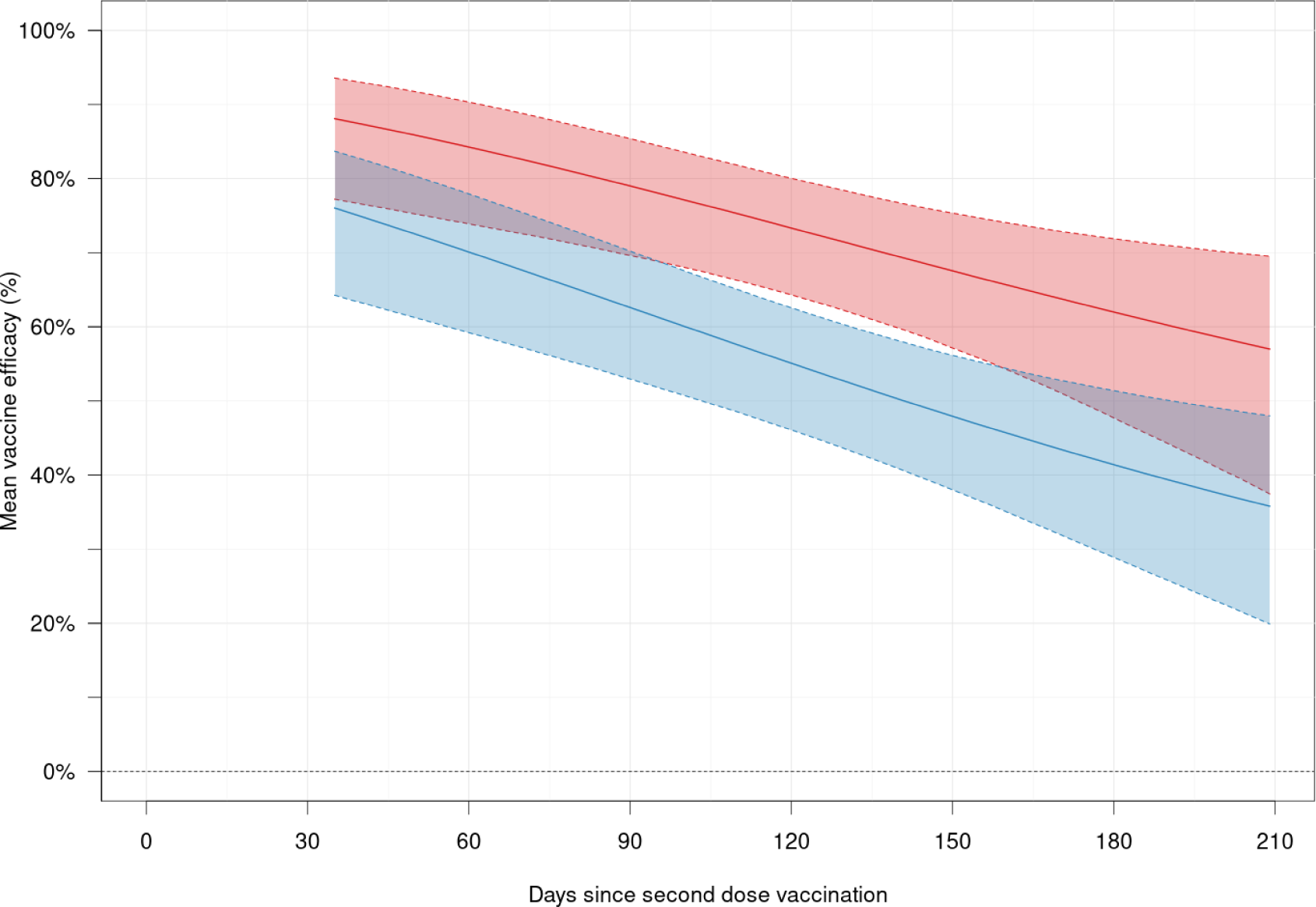
Vaccine efficacy as a function of time since second dose vaccination. The red curve and shaded region shows the posterior median and associated 95% credible interval for VE against symptomatic COVID-19 infection, and the blue curve and shaded region shows the posterior median and 95% credible interval for VE against all positive COVID-19 tests.

Levels of protection varied based on individual antibody responses. At day 28, the predicted 25%, 50% and 75% antibody quantiles were 124 (95% CrI: 117, 130), 217 (208, 227), 378 (359, 398) BAU/mL respectively (Supplementary Fig. 9). The corresponding VE quantiles at day 35 against symptomatic infection were 81.4% (95% CrI: 68, 93.7), 93.1% (76.6, 99.4), 98.8% (85.2, 100), and against any COVID-19 infection were 62.1% (49.7, 75.1), 80.5% (63.1, 92.7), 93.8% (77.1, 99.1) (Supplementary Fig. 8). At day 182, the respective antibody quantiles were 26 (23, 29), 49 (45, 54), 91 (82, 102) BAU/mL, and corresponding VE quantiles were 46.3% (16.1, 65.3), 58.8% (44.9, 69.5), 73.9% (62.6, 85.6) against symptomatic infection, and 23.6% (−0.8, 41.5), 35.5% (21.6, 47.1), 52.5% (42.1, 62.3) against any COVID-19 infection.

### Covariate effects on antibody levels and vaccine efficacy waning

We report the estimated effects of different covariates on initial antibody response, subsequent antibody decay, and vaccine efficacy over time. Our model predicts a large decrease in antibody levels for those with shorter intervals between the first and second dose (Fig. 4, Supplementary Table 5). Compared with participants with dose intervals of ≥12 weeks, participants with dose intervals of 9-11 weeks, 6-8 weeks and <6 weeks have predicted anti-spike IgG antibody levels 83% (95% CrI: 74, 93), 70% (61, 81), 51% (43, 61) as high respectively, at 28 days after the second dose. We also predict a larger antibody response due to receiving a low dose as the first dose compared with a standard dose, although 95% credible intervals contain a null effect. We predict lower antibody responses at 28 days after the second dose in older age groups, and among healthcare workers. Higher antibody responses are predicted in females than males, and in non-white participants. Shorter half-lives of antibody levels are predicted for females, and individuals with a BMI ≥ 30 (Fig. 4, Supplementary Table 5).

**Figure 4:**
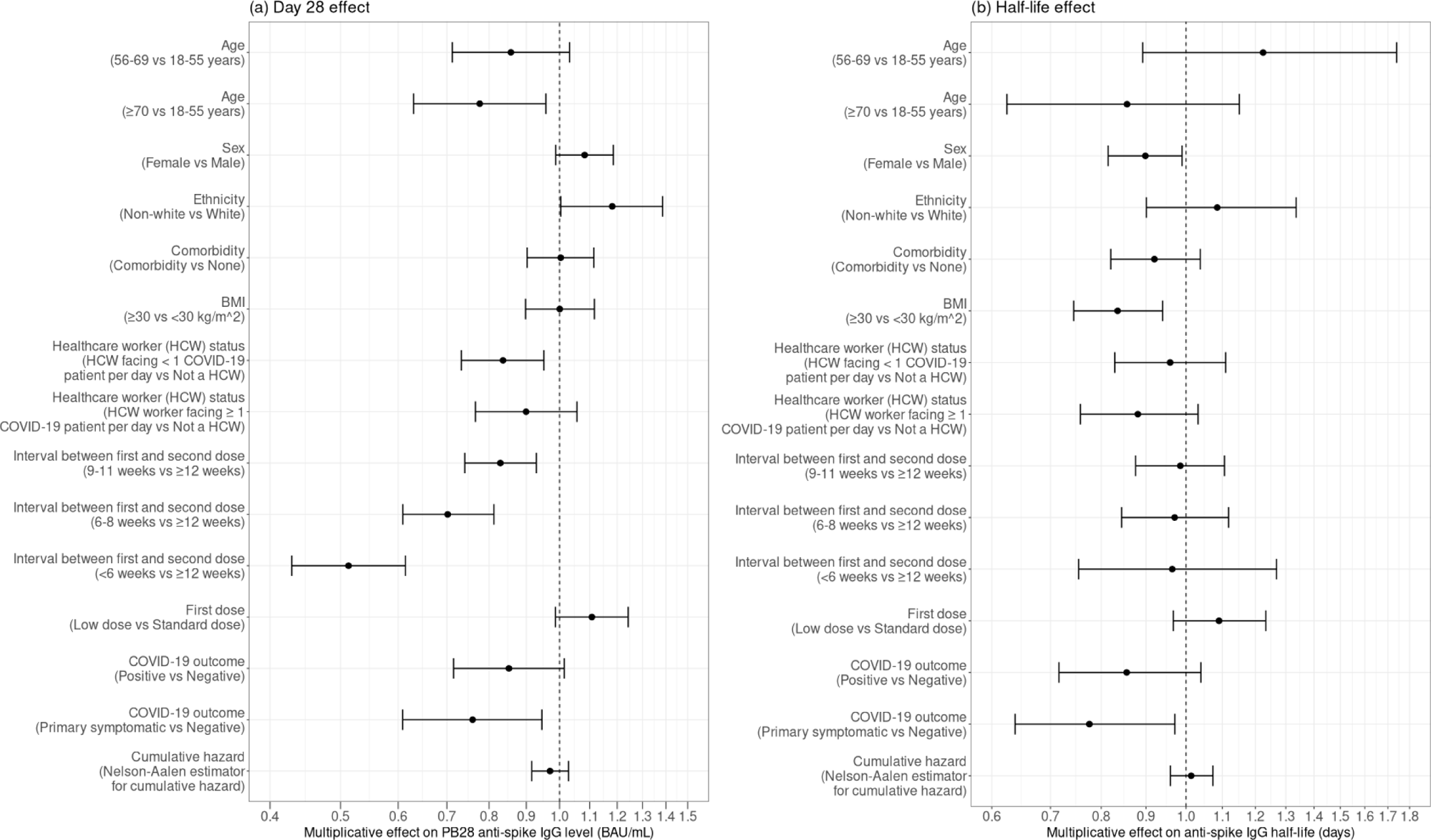
Covariate effects on anti-spike IgG antibody levels. The multiplicative effects on **(a)** day 28 anti-spike IgG antibody levels and **(b)** half-life of anti-spike IgG antibody decay for different covariates. The effects are plotted on a log scale. These results are also given in Supplementary Table 5.

To consider covariate effects on VE, we compare with reference values of age 18-55 years, female, white ethnicity, no comorbidities, BMI <30 kg/m^2^, ≥12 week interval between first and second dose, non-HCW (non-healthcare worker), and receiving a standard dose as their first dose. We then vary each of these covariates and plot VE for each covariate combination against symptomatic infection in Fig. 5 and against all COVID-19 infection in Supplementary Fig. 10. We see that shorter intervals between first and second dose predict a lower VE throughout the study. VE against symptomatic infection at day 35, 97 and 189 was 93.1% (CrI: 81.6, 97.6), 85.0% (72.1, 92.1) and 68.7% (53.2, 77.4) respectively for a reference individual (with ≥12 week interval), compared with 83.2% (71.6, 91.0), 71.7% (58.2, 80.4) and 55.0% (27.0, 69.2) respectively for an individual with <6 week interval. We also observe a lower efficacy after around 90 days in individuals aged 70 years or older compared with the 18-55 and 56-69 years age groups. VE for an individual aged 70 years or older at day 35, 97 and 189 was 90.7% (78.8, 96.4), 79.2% (67.5, 88.2) and 59.9% (35.2, 72.7) respectively, compared with a reference individual above.

**Figure 5:**
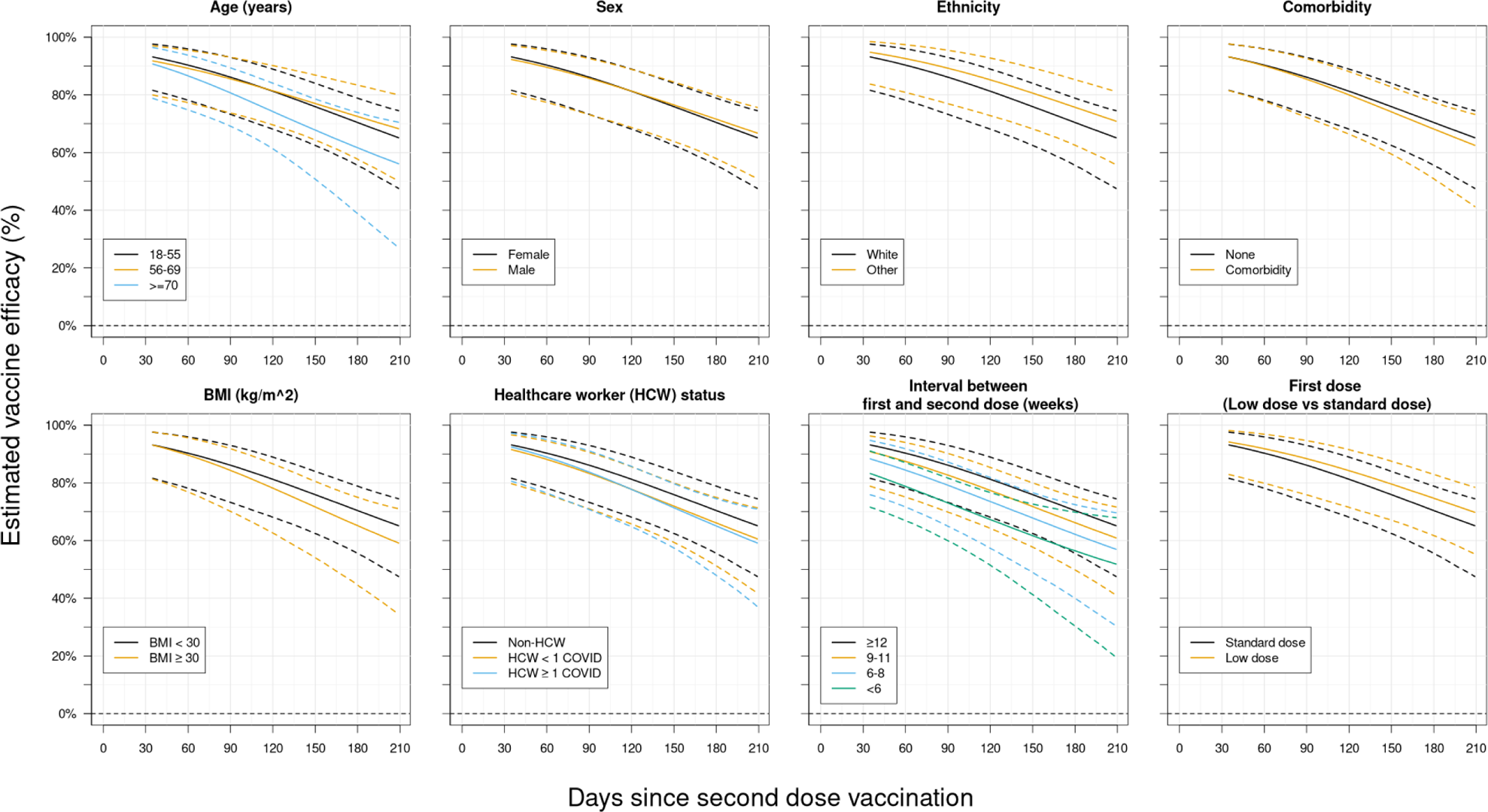
Vaccine efficacy against symptomatic COVID-19 infection plotted against time since vaccination for different covariate combinations. The black lines represent an individual with reference covariates: age 18-55, female sex, white ethnicity, no comorbidities, BMI<30 kg/m^2^, ≥12 week interval between first and second dose, non-HCW (healthcare worker), and receiving a standard dose as their first dose. All other coloured lines represent an individual with one of the above covariates changed from these reference values. The solid lines show posterior medians and dashed lines 95% credible intervals. “HCW < 1 COVID” refers to a healthcare worker facing <1 COVID-19 patient per day, “HCW ≥ 1 COVID” refers to a healthcare worker facing ≥1 COVID-19 patient per day.

## Discussion

We estimated correlates of protection, vaccine efficacy waning and antibody waning after two doses of the ChAdOx1 nCoV-19 (AZD1222) vaccine, against any COVID-19 infection and symptomatic COVID-19 infection. We fit a two-stage joint model using multiple imputation [31] which accounts for antibody waning and measurement error, allowing the risk of infection to depend on the estimated anti-spike IgG level at exposure. We found antibody levels are predictive of vaccine efficacy, which wanes over time, but continues to protect against the Alpha variant 209 days after the second dose. We estimated mean VE against symptomatic infection to be 88.1% (95% CrI: 77.2, 93.6) at day 35, waning to 60.4% (44.6, 71.0) at day 189 (Results). There was noticeable variation in VE between individuals, depending on the strength of their antibody response. We reported that a longer interval between the first and second vaccine dose gives improved protection, adding to existing evidence [34, 35, 9]. VE after 3 months since second dose vaccination was also lower in individuals aged 70 years or older than in younger age groups. We estimated “exposure-proximal” correlates of protection by relating modelled antibody levels at the time of exposure to the risk of COVID-19 infection. This approach more closely matches the biological mechanism than relating peak antibody levels to risk of infection. Table 1 compares our methods and results with previous studies of COVID-19 correlates of protection. As outlined in the introduction, previous analyses have demonstrated both binding and neutralising antibodies measured at a fixed time after vaccination are associated with VE against COVID-19, including a study analysing data from the same COV002 trial [1], an analysis of data from a different trial of the ChAdOx1 nCoV-19 vaccine [2], as well as analyses of trials of other vaccines [3, 4, 5], and booster doses [6, 7]. These studies compared peak antibody levels and risk of infection, and generally report wide confidence intervals due to a lack of power. They did not account for antibody waning between the time of measurement and exposure, which may introduce a bias[8]. Other studies have estimated correlates of protection by relating antibody levels at the time of exposure to the risk of infection, by using a recent antibody measurement [9, 10], or a model to predict antibody decay over time [11]. However the studies using a recent antibody measurement (Wei et al. 2022, 2023) used observational data from a cohort study, and did not account for antibody waning between measurement and exposure, which may introduce bias [9, 10]. The study modelling antibody levels over time (Follmann et al. 2023) assumed the rate of antibody decay to be the same for all individuals in the study, which may introduce bias if it decay rates vary [11]. All the above studies only used one antibody measurement per individual, meaning they are unable to account for measurement error in the antibody levels, which may bias the analysis [12]. None of these prior studies relate protection against COVID-19 infection to antibody levels at the time of exposure while also accounting for measurement error or variation between individual rates of antibody level decay.

**Table 1:**
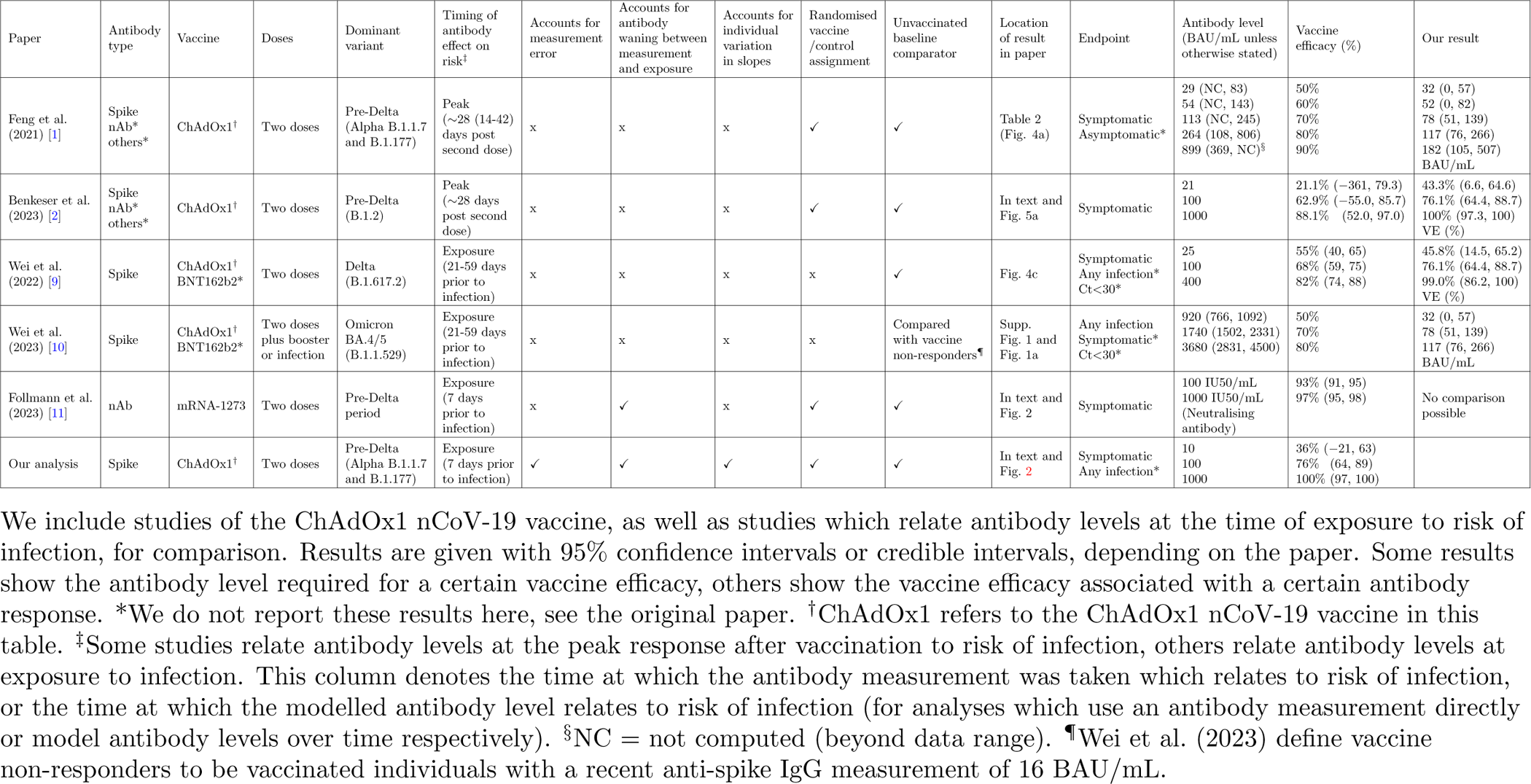
Comparison of methods and results with previous correlates of protection studies.

| Paper | Antibody type | Vaccine | Doses | Dominant variant | Timing of antibody effect on risk<sup>†</sup> | Accounts for measurement error | Accounts for antibody waning between measurement and exposure | Accounts for individual variation in slopes | Randomised vaccine /control assignment | Unvaccinated baseline comparator | Location of result in paper | Endpoint | Antibody level (BAU/mL unless otherwise stated) | Vaccine efficacy (%) | Our result |
| --- | --- | --- | --- | --- | --- | --- | --- | --- | --- | --- | --- | --- | --- | --- | --- |
| Feng et al. (2021) [4] | Spike nAb* others* | ChAdOx1<sup>†</sup> | Two doses | Pre-Delta (Alpha B.1.1.7 and B.1.177) | Peak (~28 (14-42) days post second dose) | x | x | x | ✓ | ✓ | Table 2 (Fig. 4a) | Symptomatic<br>Asymptomatic* | 29 (NC, 83)<br>54 (NC, 143)<br>113 (NC, 245)<br>264 (108, 806)<br>899 (369, NC)<sup>§</sup> | 50%<br>60%<br>70%<br>80%<br>90% | 32 (0, 57)<br>52 (0, 82)<br>78 (51, 139)<br>117 (76, 266)<br>182 (105, 507)<br>BAU/mL |
| Benkeser et al. (2023) [2] | Spike nAb* others* | ChAdOx1<sup>†</sup> | Two doses | Pre-Delta (B.1.2) | Peak (~28 days post second dose) | x | x | x | ✓ | ✓ | In text and Fig. 5a | Symptomatic | 21<br>100<br>1000 | 21.1% (-361, 79.3)<br>62.9% (-55.0, 85.7)<br>88.1% (52.0, 97.0) | 43.3% (6.6, 64.6)<br>76.1% (64.4, 88.7)<br>100% (97.3, 100)<br>VE (%) |
| Wei et al. (2022) [9] | Spike | ChAdOx1<sup>†</sup><br>BNT162b2* | Two doses | Delta (B.1.617.2) | Exposure (21-59 days prior to infection) | x | x | x | x | ✓ | Fig. 4c | Symptomatic<br>Any infection*<br>Ct<30* | 25<br>100<br>400 | 55% (40, 65)<br>68% (59, 75)<br>82% (74, 88) | 45.8% (14.5, 65.2)<br>76.1% (64.4, 88.7)<br>99.0% (86.2, 100)<br>VE (%) |
| Wei et al. (2023) [10] | Spike | ChAdOx1<sup>†</sup><br>BNT162b2* | Two doses plus booster or infection | Omicron BA.4/5 (B.1.1.529) | Exposure (21-59 days prior to infection) | x | x | x | x | Compared with vaccine non-responders*<sup>¶</sup> | Supp. Fig. 1 and Fig. 1a | Any infection<br>Symptomatic*<br>Ct<30* | 920 (766, 1092)<br>1740 (1502, 2331)<br>3680 (2831, 4500) | 50%<br>70%<br>80% | 32 (0, 57)<br>78 (51, 139)<br>117 (76, 266)<br>BAU/mL |
| Follmann et al. (2023) [11] | nAb | mRNA-1273 | Two doses | Pre-Delta period | Exposure (7 days prior to infection) | x | ✓ | x | ✓ | ✓ | In text and Fig. 2 | Symptomatic | 100 IU50/mL<br>1000 IU50/mL (Neutralising antibody) | 93% (91, 95)<br>97% (95, 98) | No comparison possible |
| Our analysis | Spike | ChAdOx1<sup>†</sup> | Two doses | Pre-Delta (Alpha B.1.1.7 and B.1.177) | Exposure (7 days prior to infection) | ✓ | ✓ | ✓ | ✓ | ✓ | In text and Fig. 2 | Symptomatic<br>Any infection* | 10<br>100<br>1000 | 36% (-21, 63)<br>76% (64, 89)<br>100% (97, 100) |  |

We include studies of the ChAdOx1 nCoV-19 vaccine, as well as studies which relate antibody levels at the time of exposure to risk of infection, for comparison. Results are given with 95% confidence intervals or credible intervals, depending on the paper. Some results show the antibody level required for a certain vaccine efficacy, others show the vaccine efficacy associated with a certain antibody response. \*We do not report these results here, see the original paper. <sup>†</sup>ChAdOx1 refers to the ChAdOx1 nCoV-19 vaccine in this table. <sup>‡</sup>Some studies relate antibody levels at the peak response after vaccination to risk of infection, others relate antibody levels at exposure to infection. This column denotes the time at which the antibody measurement was taken which relates to risk of infection, or the time at which the modelled antibody level relates to risk of infection (for analyses which use an antibody measurement directly or model antibody levels over time respectively). <sup>§</sup>NC = not computed (beyond data range). <sup>¶</sup>Wei et al. (2023) define vaccine non-responders to be vaccinated individuals with a recent anti-spike IgG measurement of 16 BAU/mL.

We used a two-stage joint model to relate modelled antibody levels at the time of exposure to the risk of infection. Joint models have been shown to reduce bias and improve power in estimating the relationship between longitudinal outcomes (e.g. antibody levels) and the time until an event (e.g. infection) [36, 25], as well as in estimating the trajectory of the longitudinal variable (antibody levels) [36]. Moreno-Bentacur et al. (2018) previously found using a multiple imputation-based joint model reduced bias, when compared with using the longitudinal observations directly to predict risk [25]. We include multiple antibody measurements per individual where available, allowing us to account for measurement error and variation between individual rates of decay. We further account for the informative censoring of antibody data after infection. We are not aware of any previous work applying joint modelling to understand how vaccine-induced immune responses relate to risk of infection for COVID-19 or any other infectious disease.

We observe higher vaccine efficacy with much lower uncertainty at the same level of antibody, compared with the studies relating peak antibody levels to risk of infection [1, 2] (Table 1). The lower efficacy these studies report is likely due to antibody waning between the measurement and exposure, meaning higher initial antibodies would be required to give the same protection at exposure. These models also do not account for measurement error, which introduces noise to the data, likely causing the observed protective effect to be diluted [12]. Our results are broadly similar to those of Wei et al. (2022) who relate antibody levels at the time of exposure to risk of infection [9] (Table 1). Much higher antibody levels are required for similar levels of protection against the Omicron variant [10]. We are unable to compare with the results of Follmann et al. (2023), as their paper uses neutralising antibodies, whereas we use anti-spike IgG [11].

We estimated the peak antibody response at 28 days after ChAdOx1 nCoV-19 vaccination, the subsequent rate of decay, and associated covariate effects. Table 2 compares our methods and results with previous studies. Wei et al. (2022) previously used a similar approach applied to non-randomised cohort data [9]. Their model does not account for the aforementioned informative censoring of antibody measurements, which may lead to bias [36]. This is because individuals with lower antibody levels will be more likely to test positive for COVID-19, after which antibody measurements will be excluded from analysis. Thus individuals with lower antibody levels are less likely to have antibody measurements available, introducing informative missing data, which should be accounted for. However, as cases are rare in the population, this bias may be small. We estimated higher peak anti-spike IgG levels than Wei et al. study [9] (Table 2). This may be partly due to differences in the populations enrolled in the two studies. Our estimated half-life of antibody decay was broadly similar to previous results (Table 2). We observed individuals with longer dose intervals had higher peak antibody levels (Fig. 4, Supplementary Table 5), as previous studies have reported [35, 34, 9]. We also observed higher peak antibody levels in younger individuals (18-55 vs ≥70), non-healthcare workers and non-white participants. The higher peak in non-white participants was also observed by Wei et al. (2022) [9], however there is no consensus on the effect of age and healthcare work on antibody levels (Table 2). We reported slower antibody decay in participants with a BMI<30kg/m^2^ (Fig. 4, Supplementary Table 5), and detected slightly slower antibody decay in males, however this does not translate to a noticeable difference in VE waning (Fig. 5, Supplementary Fig. 10). Neither of these differences have been reported elsewhere and it is unclear if they represent true effects. We did not detect the other effects on peak antibody observed by Wei et al. (2022) [9].

**Table 2:** Comparison of methods and results with previous studies of antibody level waning.

| Paper | Vaccine | Randomised vaccine /control assignment | Account for informative censoring at infection | Random individual slopes | Antibody type | Time of reported peak antibodies | Two-dose /booster /prior infection | Peak anti-spike IgG antibody level (BAU/mL) | Antibody half-life (days) | Covariates related to increased peak antibody levels | Covariates related to slower antibody decay |
| --- | --- | --- | --- | --- | --- | --- | --- | --- | --- | --- | --- |
| Wei et al. (2022) [9] | ChAdOx1 nCoV-19 BNT162b2* | x | x | ✓ | Anti-spike IgG | 21 days post second dose | Two-dose | 184 (183, 185) | Mean 79 (78, 80) | Older age<br>Female<br>Non-white<br>No LTHC<br>Prior infection<br>Healthcare worker<br>Longer dose interval<br>More deprivation | Younger age<br>White<br>No LTHC |
| Aldridge et al. (2022) [41] | ChAdOx1 nCoV-19 BNT162b2* | x | x | x | Anti-spike IgG | 28 days post second dose | Two-dose | - | 105.9 | None | None |
| Wei et al. (2023) [10] | ChAdOx1 nCoV-19 BNT162b2 | x | x | ✓ | Anti-spike IgG | 21 days post second dose | Booster: BNT162b2 mRNA-1273 Infection | - | 78 (72, 86)<br>63 (61, 66)<br>96 (80, 119) | Younger age<br>Longer dose interval | Not reported |
| Our analysis | ChAdOx1 nCoV-19 | ✓ | ✓ | ✓ | Anti-spike IgG | 28 days post second dose | Two-dose | 217 (208, 227) | Median 74 (69, 79) | Younger age (18-69)<br>Non-white<br>Longer dose interval<br>Not HCW | Male<br>BMI <30 kg/m<sup>2</sup> |

We include studies of two doses of the ChAdOx1 nCoV-19 vaccine, and studies after a subsequent booster vaccine, focussing on anti-spike IgG antibody waning. Results are given with 95% confidence intervals or 95% credible intervals, depending on the paper. We report covariate effects where the null effect is not included in the 95% interval. HCW = healthcare worker, LTHC = long-term health condition. \*We do not report these results here, see the original paper.

We reported predicted vaccine efficacy over time since second dose from our model, calculated from randomised controlled trial data. One previous analysis considered waning after a first dose of ChAdOx1 nCoV-19 [34], however the analysis was underpowered. Apart from this, we are not aware of any studies of ChAdOx1 nCoV-19 vaccine efficacy waning evaluated on randomised data. Table 3 and Supplementary Table 8 compare our methods and results to previous studies of waning vaccine efficacy, with a focus on studies of the ChAdOx1 nCoV-19 vaccine. These studies all use non-randomised observational data, including test-negative design studies [13, 14, 15, 18], and cohort studies [37, 15, 16, 17]. The test-negative design may introduce bias due to unmeasured differences in health-seeking behaviour between the vaccinated and unvaccinated arms, likely reducing the observed VE [20, 21]. Results may also not carry over to the wider population, as those included in the study will likely test more frequently. Cohort studies are similarly vulnerable to biases due to unmeasured confounding causing differences in risk of infection between the unvaccinated and vaccinated populations. Such biases may increase over time, as the unvaccinated group are increasingly unvaccinated by choice, which may correlate with fewer health-seeking behaviours. Further, accumulation of more undetected infections in the unvaccinated arm may lead to VE waning to appear greater, due to the so-called depletion of susceptibles bias [38]. This bias may occur in randomised or observational data. Asymptomatic swabbing may negate this bias by increasing the proportion of infections which are detected. Rates of asymptomatic swabbing were high in our study [39], with around 75% of participants returning an asymptomatic test on a given week. Rates of asymptomatic swabbing were likely much lower in the other studies, where participants were not encouraged to swab weekly.

**Table 3:** Comparison of methods and results with previous vaccine efficacy waning studies, against the Alpha and Delta variants.

| Paper | Primary course vaccine | Dominant variant | Two-dose /booster /prior infection | Study design | Randomised vaccine /control assignment | Baseline comparator | Outcome | Location of result in paper | Group | Weeks since dose | Vaccine efficacy (%) |
| --- | --- | --- | --- | --- | --- | --- | --- | --- | --- | --- | --- |
| Our analysis<sup>†</sup> | ChAdOx1 nCoV-19 | Alpha | Two dose | Randomised Controlled Trial | ✓ | Unvaccinated | Symptomatic | Fig. 5 | Age 18-55<br>Age 56-69<br>Age 70+ | 5<br>10<br>20<br>26<br>5<br>10<br>20<br>26<br>5<br>10<br>20<br>26 | 93.1 (81.6, 97.6)<br>89.0 (76.6, 95.1)<br>77.7 (64.5, 85.7)<br>70.0 (55.0, 78.5)<br>91.8 (80.0, 97.0)<br>88.1 (76.1, 94.7)<br>78.5 (66.3, 88.0)<br>72.2 (57.2, 83.1)<br>90.7 (78.8, 96.4)<br>84.7 (73.0, 92.4)<br>69.9 (54.5, 80.4)<br>61.3 (38.1, 73.6) |
|  |  |  |  |  |  |  | Any infection | Supp. Fig. 10 | Age 18-55<br>Age 56-69<br>Age 70+ | 5<br>10<br>20<br>26<br>5<br>10<br>20<br>26<br>5<br>10<br>20<br>26 | 84.3 (70.6, 91.8)<br>77.2 (62.7, 86.1)<br>60.5 (46.1, 70.1)<br>50.6 (34.8, 60.1)<br>81.8 (68.3, 90.4)<br>75.6 (62.4, 85.3)<br>61.6 (48.0, 73.7)<br>53.4 (37.0, 66.5)<br>79.9 (66.4, 89.0)<br>70.4 (57.6, 80.9)<br>50.4 (34.2, 62.8)<br>40.1 (18.4, 53.8) |
| Andrews et al. (2022a) [13] | ChAdOx1 nCoV-19 BNT162b2* | Alpha Delta* | Two dose | Test-negative | x | Unvaccinated | Symptomatic | Table S11 | Age 16+<br>Age 40-64<br>Age 65+ | 2-9<br>10+<br>2-9<br>2-9 | 82.4 (79.6, 84.7)<br>76.2 (49.8, 88.7)<br>81.7 (76.0, 86.1)<br>88.2 (82.2, 92.1) |
| Nordström et al. (2022) [37] | ChAdOx1 nCoV-19 BNT162b2* mRNA-1273* | Alpha and Delta (Combined) | Two dose | Cohort | x | Unvaccinated | Any inection | Table 2 | Alpha, Delta and all ages combined | 2-4<br>4-9<br>9-17<br>≥ 17 | 68 (58, 70)<br>49 (28, 64)<br>41 (29, 51)<br>-19 (-98, 28) |
| Skowronski et al. (2022) [14] | ChAdOx1 nCoV-19 BNT162b2* mRNA-1273* Mixed schedules* | Alpha Delta | Two dose | Test-negative | x | Unvaccinated | Any inection Hospitalisation* | Supp. Table 12 | Quebec (Alpha and Delta)<br>British Columbia (Delta) | 4-7<br>8-11<br>20-23<br>24-27<br>4-7<br>8-11<br>20-23<br>24-27 | 90 (84, 94)<br>85 (81, 88)<br>70 (64, 75)<br>56 (43, 65)<br>77 (71, 82)<br>76 (73, 79)<br>74 (70, 78)<br>67 (48, 80) |

We include studies of two doses of the ChAdOx1 nCoV-19 vaccine, prior to the Omicron variant. Results are given with 95% confidence intervals or 95% credible intervals, depending on the paper. Results are presented for different age groups, where appropriate. \*We do not report these results here, see the original paper. <sup>†</sup>We report here the predicted vaccine efficacy for a reference individual in the given age group, with covariates: female, white, no comorbidities, BMI<30 kg/m<sup>2</sup>, ≥12 week interval, not a healthcare worker, standard dose.

Our analysis has some advantages over these methods. The data comes from a randomised single-blinded controlled trial, avoiding the potential biases of an observational cohort study and a test-negative case-control study. We model antibody waning and vaccine efficacy waning jointly over time, which may improve the efficiency of our estimates [22]. Further, the length of follow-up in our study after second dose was relatively long at over 29 weeks. However, the sample size of our data is much smaller than most of these studies, hence we report wider CrIs. Table 3 and Supplementary Table 8 compare our methods and results to previous studies of vaccine efficacy waning against different variants. Results from previous studies varied by variant, with lower efficacy generally predicted against Delta than Alpha, and lower still against Omicron (Table 3, Supplementary Table 8). Our study reports efficacy over time since second dose against the Alpha variant. Note the predicted vaccine efficacy for a reference individual reported in Table 3 and Figure 5 was slightly higher than the mean vaccine efficacy across all individuals (Fig. 3), primarily because the mean vaccine efficacy averaged over all individuals including those who had a shorter dose interval than 12 weeks. We predicted a slightly higher vaccine efficacy against symptomatic infection with the Alpha variant after two doses of ChAdOx1 nCoV-19 (Table 3, Fig. 5) than Andrews et al. (2022a) in all age groups, although confidence intervals overlapped [13]. Other studies were conducted at least partly in periods where the Delta or Omicron variants were dominant, and generally reported lower vaccine efficacy. Reported efficacy varied noticeably between studies (Table 3, Supplementary Table 8). Differences in results between studies may be due to different proportions of circulating variants, rates of asymptomatic swabbing, and characteristics of the study populations, as well as the depletion of susceptibles bias, and previously mentioned biases due to unmeasured confounding. Our results against the Alpha variant, calculated on data from a randomised single-blinded study, are free from bias due to unmeasured confounding.

Our data was collected in a period when the Alpha variant was dominant, and considered individuals without prior infection who received two doses of vaccine. In contrast, the Omicron variant is currently dominant, and many individuals have received three or more doses of vaccine and been previously infected. Therefore our results may not be directly applicable to current and future situation. To aid with this, we performed a supplementary analysis extrapolating our results to examine what the efficacy over time since second dose might have been, in the hypothetical scenario where the study was conducted in the Omicron BA.4/5 period. The analysis suggests a weak initial protection against Omicron BA.4/5, waning to negligible protection after 6 months. See the Supplementary Information (Sections 1.4.6, 2.1, 3, Supplementary Fig. 11) for further details.

We further reported covariate effects on vaccine efficacy, although this analysis lacked power. We observed lower vaccine efficacy for individuals with shorter intervals between the second dose; results from previous studies have provided varying degrees of evidence for this [40, 14, 13]. We reported lower VE in older individuals after around 3 months since the second dose (Fig. 5, Supplementary Fig. 10), with overlapping credible intervals. Faster waning [13] and lower VE [37, 16] have been previously observed in older individuals. However, these effects were not observed in all groups in the Andrews et al. study, with non-significant effects in the opposite direction observed in some groups [13], and the Nordströom et al. age estimates were unadjusted and likely confounded due to different vaccines being given to different age groups [37]. Another observational study observed higher efficacy in individuals aged 80+ years than 50-79, while suggesting this could be confounded by clinical risk [40]. Hence further research on the effect of age on vaccine efficacy is necessary. We did not observe a difference in vaccine efficacy for individuals with comorbidites, although some prior studies suggest a lower VE in clinically vulnerable individuals [16, 37, 13].

Our study has several limitions. Our results may not be directly applicable to current and future populations, for four reasons. Firstly dominant variants have changed from B.1.177 and Alpha (B.1.1.7) variants when our data was collected, to the Omicron variant today. Secondly individuals in the study were SARS-CoV-2 naïve, results will likely differ in individuals with prior infection [10]. Thirdly individuals in the study received two doses of ChAdOx1 nCoV-19, whereas three or more doses of different vaccines have been given in many populations.

Fourthly the study population differs from the general population, being predominantly white, 18-55 years old and including mostly healthcare workers. For these reasons our results are unlikely to transfer directly to the general population today, although qualitative aspects of our results, and our methods, remain relevant. We further only consider the effect of anti-spike IgG binding antibodies, and do not measure neutralising antibodies or cellular immune responses. It may be that the protection conferred by other immune responses depends on the antibody level, or changes with time, whereas we assumed a constant effect. We did not consider the effect on risk of hospitalisation, severe disease and death due to insufficient data. Fewer antibody observations were available at the later timepoints (PB90 and PB182 study visits), causing greater uncertainty in our estimates at later times. We assumed a 7 day gap from exposure to reported infection; deviation from this assumption may introduce a small bias to our analysis. We did not account for the change in dominant variant from B.1.177 to B.1.1.7 (Alpha) during the study, however we expect the impact of this on VE to be negligible [42].

The dose of virus that individuals were exposed to may have varied throughout the study, as social distancing policies changed. Further, viral load is likely to have varied between individuals depending on previous exposure to COVID-19, and the variant to which they were exposed. We have not accounted for differences in viral load or dose of virus in this analysis. In summary, we report correlates of protection against any COVID-19 infection and symptomatic infection from a randomised COVID-19 vaccine study, using binding antibody responses at the time of exposure. We further report vaccine efficacy waning over time, antibody waning over time, and associated covariate effects. We use a joint model which accounts for antibody measurement error, the waning of antibody levels over time, and censoring of antibody observations at COVID-19 infection. This may reduce bias and increase power in the analysis compared with previous approaches.

## Methods

### Study description

We analysed data from COV002 (registration NCT04400838), a phase 2/3 randomized single-blind vaccine efficacy (VE) trial conducted across 19 sites in the United Kingdom. A full description of the trial including immunogenicity, efficacy, and safety data, and the protocol has been previously published [32, 33, 34, 42]. An analysis of immune correlates of protection, relating antibody measurements 28 days after the second dose with protection against infection, was previously published [1].

This study was approved in the United Kingdom by the Medicines and Healthcare products Regulatory Agency (MHRA), reference 21584/0428/001–0001, and the South-Central Berkshire Research Ethics Committee, reference 20/SC/0179. All participants provided informed consent. Participants in the study were randomized to receive ChAdOx1 nCoV-19 (AZD1222) or a MenACWY control vaccine. The randomization ratio (ChAdOx1 nCoV-19:MenACWY) differed by study cohort, and was either 1:1, 5:1, or 3:1. Open label groups are not included in this analysis.

### Study endpoints and outcomes

Participants were sent weekly reminders to contact their study site if they experienced any of the primary symptoms of COVID-19 (fever ≥37.8 °C; cough; shortness of breath; anosmia or ageusia) and were then assessed in clinic, by taking a nose and throat swab by a nucleic acid amplification test (NAAT). In addition, participants were asked to complete a nose and throat swab at home each week.

The outcomes for this analysis were (1) primary symptomatic COVID-19 infection, that is a nucleic acid amplification test positive (NAAT+) swab with at least one qualifying symptom, (2) any COVID-19 infection, that is any NAAT+ swab.

All endpoints were evaluated by a blinded independent clinical review committee. The date of onset of infection was determined by the committee, as the earliest time at which there was evidence of infection. Evidence included NAAT+ swabs and self-reported symptoms based on telephone contact and study visits.

### Antibody measurements

A proportion of serum samples from vaccine recipients at the PB28, PB90 and PB182 study visits were tested for anti-SARS-CoV-2 Spike IgG in a single laboratory assay. For PB28 samples, all NAAT+ cases were tested if sample volume allowed, and a proportion of non-cases were tested. Samples were tested blinded to case status. The data from noncases was obtained first, and consisted mainly of the samples processed for the initial application for emergency use, which needed 15% of samples included in the efficacy cohort to be processed on validated assays. Subsequent to this PB28 samples from NAAT+ cases were sent for testing as they occurred, if not already including the 15%. A proportion of those participants with samples tested at PB28 were additionally tested at either PB90 only, or PB90 and PB182. We assume the samples to be missing at random, i.e. the missingness depended only on factors observed in the study. To account for the missing data, factors associated with sample availability were included as covariates in the model for antibody decay (see ‘Antibody decay model’ below).

Anti-SARS-CoV-2 Spike IgG was measured by a multiplex immunoassay on the MSD platform at PPD Laboratories. The assay sequences were based on the ancestral sequences from Wuhan, China. Assay validation included precision and ruggedness, dilutional linearity, selectivity, and relative accuracy for each SARS-CoV-2 antigens. Post-validation studies for stability and for conversion to the WHO standard, as well as the establishment of a cut-point, were performed. The lower limit of quantification (LLOQ) for anti-spike IgG was 33 AU/mL (0.21 BAU/mL). We excluded 8 anti-spike IgG results as outliers - three results at PB28 and four at PB90 whose anti-spike IgG levels were especially low, and one results at PB90 whose levels were especially high (Supplementary Table 3, Supplementary Fig. 2). The individuals from whom these samples were taken, and any other antibody measurements are included in the analysis - only the outlying antibody measurements are excluded. Outliers were excluded as they may bias the results [43].

The assay was analysed in its original scale. Results were then converted to the WHO international standard units, binding antibody units per mL (BAU/mL), by multiplying by a conversion factor supplied by the laboratory. The anti-spike IgG conversion factor from arbitrary units per mL (AU/mL) to WHO standard binding antibody units per mL (BAU/mL) was 0.00645 (95% confidence interval (CI): 0.00594, 0.00701). We did not apply the CI, as it represents the uncertainty due to measurement error in the assay, and our antibody model already accounts for this.

### Study design

We included a subset of participants in the COV002 trial in our study, who met the following eligibility criteria: received two doses of ChAdOx1 nCoV-19 or MenACWY control vaccine, baseline seronegative to the SARS-CoV-2 N protein at first vaccination, and followed up to at least day 21 post second vaccination with no evidence of prior infection. Vaccinated participants received a first and second dose: either two standard doses, low dose followed by standard dose, or two low doses. Nine participants who received mixed schedules in error (one dose of ChAdOx1 nCoV-19 and one dose of MenACWY control) were excluded from the analysis (Supplementary Fig. 1). One control individual had missing BMI, we imputed this with the mean value in the study.

Participants were considered at-risk of the study event from day 21 post second dose. The at-risk period ended at the first of: the onset of COVID-19 infection, withdrawal, unblinding, or the analysis cut-off date 30 June 2021. Antibody levels were assumed to relate to risk of infection 7 days later, to account for the gap between first being exposed to COVID-19 until a case is reported in the trial. Thus antibody measurements are excluded if taken prior to 14 days after the second dose, after the end of the at-risk period, or less than 7 days before the end of the at-risk period.

Participants who tested NAAT+ during the at-risk period were defined as cases, those who also had at least one qualifying symptom were defined as symptomatic cases, while those who did not return a positive test during the at-risk period were defined as non-cases.

### Statistical Analysis

We modelled antibody decay and the risk of COVID-19 infection in a two-stage joint model for longitudinal and survival data. Joint models are a popular method to consider the relationship between a longitudinal biomarker and the risk of a health outcome, especially in studies of disease progression such as HIV and cancer trials [8]. We employed a two-stage multiple imputation approach to overcome convergence issues that arose when attempting to fit a full joint model. A mathematical formulation of the model, and an explanation of how the two-stage method approximates a Bayesian joint model, are given in the Supplementary Methods.

#### Antibody decay model

In the first stage, we fit a log-linear Bayesian mixed effects model to the antibody decay after vaccination for ChAdOx1 nCoV-19 recipients (see Supplementary Methods). The Bayesian approach easily accounts for the complex structure of the data and the model, including missing antibody observations for some individuals, and unknown true antibody levels which change over time. We first standardised the data; after running the model we transformed the parameters back to the scale of the original dataset. The log antibody observations were assumed to have t-distributed zero-mean random error, with 4 degrees of freedom and unknown scale (Supplementary Methods). The latent true log antibody values were assumed to decay linearly from 14 days post second dose. The model included an effect on both the peak antibody response (intercept) and rate of decay (slope) due to the following covariates: age group (18-55 years, 56-69 years, 70+ years), sex (female, male), ethnicity (white, non-white), comorbidity (none, comorbidity), BMI (<30 kg/m^2^, ≥30), time interval between first and second dose (≥12 weeks, 9-11 weeks, 6-8 weeks, <6 weeks), healthcare worker (HCW) status (not a HCW, HCW facing <1 COVID-19 patient per day, HCW facing ≥1 COVID-19 patient per day), initial dose (standard dose, low dose). The probability of missing antibody data was observed to be related to whether an individual tested positive and their follow-up time. To account for this, we also included covariate effects on the intercept and slope due to: (i) returning a positive test during the study, (ii) being primary symptomatic, and (iii) the Nelson–Aalen estimate of cumulative hazard for any COVID-19 infection in the vaccine group, as previously suggested [44]. We allowed a random intercept and slope for each individual, as well as a correlation between the random effects. The slopes were required to be negative, by applying the negative of the exponential function to the slope parameter. This was done to avoid convergence issues in the risk of infection model due to exponentially increasing antibody levels. The model predicts antibody trajectories for all vaccinated individuals, with appropriate uncertainty. The model was fit using Hamiltonian Monte Carlo in Stan [45]. Four chains were run, each of 20,000 iterations. The first 5,000 iterations were discarded as burn-in. Convergence was checked by ensuring the Rhat convergence diagnostic (potential scale reduction factor) was less than 1.01 [46]. See the Supplementary Information (Section 2.3) for the reported computation time.

#### Risk of infection model

In the second stage, we fit a Cox model to the time until COVID-19 onset (either primary symptomatic COVID-19 infection or any COVID-19 infection), using the output from the antibody model to predict the risk of infection. We employed multiple imputation [31], which accounts for the uncertainty in the predicted antibody levels in our estimation of the hazard (instantaneous risk) for infection (see Supplementary Methods). A previously published multiple imputation-based joint model was shown to reduce bias compared with single imputation “naïve” two-stage approaches [25]. We first sampled 60,000 antibody trajectories from the first stage for each vaccinated individual, from which we imputed the predicted antibody levels at each relevant time. For each sample, we fit a Cox model for the time to infection. The baseline hazard varies with calendar time, to account for the changing rates of COVID-19 in the population during the pandemic, and is stratified by study site, to account for different rates across geographic regions. The hazard for a given vaccinated individual *i* at time *t* then depends on the effect *γ* of their imputed antibody levels A*_i_*(*t −* 7) at time *t −* 7, and a direct effect due to vaccination *ζ*, unrelated to antibody levels. We further included covariate effects which may affect the risk of infection: age group (18-55 years, 56-69 years, 70+ years), sex (female, male), ethnicity (white, non-white), comorbidity (none, comorbidity), BMI (<30 kg/m^2^, ≥30), healthcare worker (HCW) status (not a HCW, HCW facing <1 COVID-19 patient per day, HCW facing ≥1 COVID-19 patient per day). The hazard for a given control individual *i* at time *t* depends only on the baseline hazard and covariates, we assume no antibody or vaccine effect for control individuals.

For each Cox model, we then drew from the asymptotic distribution of the maximum likelihood estimators, to approximate a Bayesian posterior with diffuse improper priors [47]. We combined this draw with the corresponding antibody model individual intercepts and slopes to form a draw from the approximate joint distribution. Quantities of interest, such as vaccine efficacy, can then be calculated from each draw. Posterior medians and 95% CrIs are calculated based on quantiles of the posterior samples.

#### Calculating vaccine efficacy

Vaccine efficacy (VE) at a given antibody level A is then calculated as 1 minus the hazard ratio between a vaccinated individual with that level of antibody, and a control individual with matching covariates.

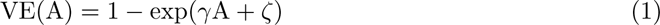

with *γ* the antibody effect and *ζ* the direct effect due to vaccination. Antibody level at a given VE is defined by the inverse of Eq (1). For individuals *i* = 1, …, *n* in the vaccine arm, let A*_i_*(*t −* 7) be the latent antibody level at time *t −* 7. Then the mean VE at time *t* is the mean VE among all the latent antibody levels of vaccinated individuals at time *t −* 7.

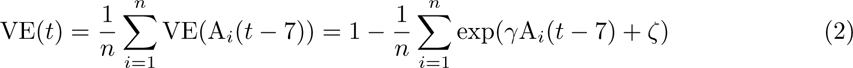

The *q*th quantile of VE at time *t*, VE*_q_*(*t*), is equal to the VE at the *q*th antibody quantile *A_q_*(*t*), VE*_q_*(*t*) = 1 − exp(*γA_q_*(*t*) + *ζ*).

#### Calculating covariate effects on vaccine efficacy

To calculate covariate effects on vaccine efficacy, we predicted efficacy in new individuals with given covariate values. Note the event indicators and Nelson–Aalen (N–A) estimate of cumulative hazard for a new individual are unknown, as they depend on the infection outcome. We first built an imputation model to sequentially impute the missing event indicators (infection, symptomatic) and N–A estimate, which appear in the longitudinal antibody model, given the known covariates. We imputed these missing covariates *n* times. For each set of imputed covariates, we drew from the predictive distribution of antibody trajectories for a new individual with the given covariate values, A^pred^(*t*), *j* = 1, …, *n*. We then averaged these to calculate the mean risk of infection given the known covariate values

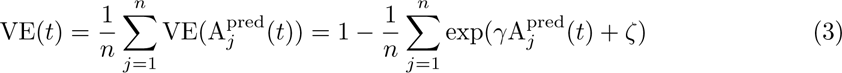

#### Software

Data analysis was performed in R version 4.3.1 [48], using RStudio [49], and relying extensively on the following packages: survival [50], RStan [51], snowfall [52], ggplot2 [53]. RColorBrewer [54] and tidyr [55] were also used. Hamiltonian Monte Carlo algorithms were run in Stan [45] via the R package RStan [51].

## Supporting information

Data for Figure 2

Data for Figure 3

Data for Figure 5

Data for Supplementary Figure 3

Data for Supplementary Figure 4

Data for Supplementary Figure 5

Data for Supplementary Figure 6

Data for Supplementary Figure 8

Data for Supplementary Figure 9

Data for Supplementary Figure 11

## Data availability

Anonymized data will be made available via the Vivli platform https://vivli.org/. Source data for the figures are provided with this paper where possible.

## Code availability

The code used for the analysis is available at GitHub repository https://github.com/danphillipsstats/COVID-joint-model-CoP.

## Acknowledgements

We thank Geoff Nicholls for a helpful discussion suggesting the use of Bayesian cut posteriors, which led us to developing the Bayesian multiple imputation method.

## Author contributions

All authors contributed to the conceptualisation of the study. SF, AJP and MV contributed to data collection. DJP, MDC and DS developed the statistical methods. DJP conducted the statistical analysis. The project was jointly supervised by MDC and DS. The first draft of the report was written by DJP and reviewed by MDC and DS. All authors critically reviewed and approved the final version.

## Ethics approval

The COV002 study was approved in the United Kingdom by the Medicines and Healthcare products Regulatory Agency (MHRA), reference 21584/0428/001 0001, and the South-Central Berkshire Research Ethics Committee, reference 20/SC/0179. All participants provided informed consent.

