## Supplementary Information for "Improved estimates of COVID-19 correlates of protection, antibody decay and vaccine efficacy waning: a joint modelling approach"

July 2, 2024

**3 Supplementary Discussion 9**

**4 Supplementary Tables** 10

**5 Supplementary Figures** 18

5 **The multiplicative covariate effects on anti-spike IgG level 28 days post second dose, and half-life of the subsequent decay in anti-spike IgG levels** . 14
6 **The multiplicative covariate effects on hazard against symptomatic COVID-19 infection and any COVID-19 infection** . . . . . . . . . . . . . . . . . . . . . . . 15
7 **Anti-spike IgG antibody levels required to give different levels of vaccine efficacy against any COVID-19 infection or symptomatic COVID-19** . . . . . 16
8 **Comparison of methods and results with previous studies considering waning vaccine efficacy against the Delta, Gamma and Omicron variants** . . . . 17

### List of Figures

1 Flow chart showing inclusion of COV002 participants in analysis cohort . . . . . . . . 18
2 Observed anti-spike IgG antibody levels over time since vaccination, including outliers and measurements from control participants . . . . . . . . . . . . . . . . . . . . . . . . . 19
3 The estimated baseline hazard of infection in the trial by calendar days . . . . . . . . 20
4 **The number of individuals at risk over time since second dose in the vaccine and control arms.** The upper green area denotes the number of ChAdOx1 nCoV-19 (AZD1222) vaccinated participants at risk over time since second dose. The lower blue area denotes the number of control participants at risk over time since second dose. As such, the upper black line denotes the total number of participants at risk over time since second dose, and the lower black line the number of control participants at risk over time. Participants are censored from the analysis upon leaving the study, returning a positive COVID-19 test, and receiving a subsequent vaccination, among other reasons. 21

5 **Histogram of the timings of anti-spike IgG samples since second dose vaccination.** Only anti-spike IgG samples from ChAdOx1 nCoV-19 (AZD1222) vaccinated participants are included. Samples from individuals more than 7 days prior to entering the at-risk period or less than 7 days before leaving the at-risk period are excluded. Hence, samples less than 7 days before returning a positive COVID-19 test, or after returning a positive test, are excluded. . . . . . . . . . . . . . . . . . . . . . . . . . . . . 22
6 **Histogram of the onset time since second dose of (a) any COVID-19 infection, (b) primary symptomatic COVID-19 infection.** (a) The blue histogram denotes the timings of the onset of any COVID-19 infection. (b) The red histogram denotes the timings of the onset of primary symptomatic COVID-19 infection. . . . . 23
7 **Covariate effects on hazard of COVID-19 infection.** The multiplicative effects due to different covariates on hazard of symptomatic COVID-19 infection and any COVID-19 infection. Points denote posterior medians and lines denote 95% credible intervals. Red denotes hazard against symptomatic infection and blue denotes hazard against any COVID-19 infection. These results are also given in Supplementary Table 6 24
8 **Quantiles of vaccine efficacy as a function of time since vaccination.** The curves show posterior medians and shaded regions 95% credible intervals. The 95%, 75%, 50%, 25% and 5% quantiles are plotted. (a) shows VE against symptomatic COVID-19 and (b) against all COVID-19 infection. . . . . . . . . . . . . . . . . . . . . . . 25
9 **Quantiles of anti-spike IgG antibody levels as a function of time since vaccination.** The curves show posterior medians and shaded regions 95% credible intervals. The 95%, 75%, 50%, 25% and 5% quantiles are plotted. . . . . . . . . . . . . . . . . . . 26
10 **Vaccine efficacy against any COVID-19 infection plotted against time since vaccination for different covariate combinations.** The black lines represent an individual with "baseline" covariates: age 18-55, female, white ethnicity, no comorbidities, BMI$<$30 kg/m$^2$, $>$12 week interval between first and second dose, non-HCW, and receiving a standard dose as their first dose. All other lines represent an individual with one of the above covariates changed from these baseline values. The solid lines show posterior medians and dashed lines 95% credible intervals. "HCW $<$ 1 COVID" refers to a healthcare worker facing $<$1 COVID-19 patient per day, "HCW $\geq$ 1 COVID" refers to a healthcare worker facing $\geq$1 COVID-19 patient per day. . . . . . . . . . . . 27
11 **Predicted relative efficacy against SARS-CoV-2 Omicron BA.4/5 variant infection as a function of time since vaccination.** The black, blue and red lines represent estimates calculated using the reported posterior mean, and upper and lower 95% credible bounds for the relationship between relative efficacy and anti-spike IgG antibody levels from Wei et al. (2023). The solid lines show posterior medians and dashed lines 95% credible intervals, for the mean relative efficacy in the study. . . . . 28

12 **Estimated true anti-spike IgG levels for 12 randomly chosen individuals.** We randomly chose 3 individuals with 0, 1, 2 and 3 antibody observations available respectively, and plotted their estimated true anti-spike IgG levels over time. The solid line represents the posterior median, and the dashed lines represent a 95% credible interval. The blue points show the observed antibody levels. To be clear, the 3 plots on each row are 3 separate individuals (not the same individual with data consecutively being added). . . . . . . . . . . . . . . . . . . . . . . . . . . . . . . . . . . . . . . . 29

### 1 Supplementary Methods

#### 1.1 Model

##### 1.1.1 Longitudinal component

Assume there are $N$ individuals in the study, of whom $n$ received two doses of the ChAdOx1 nCoV-19 vaccine (the vaccine arm), and $N-n$ received two doses of a MenACWY control vaccine (the control arm). Without loss of generality, let the dataset be ordered such that individuals $i = 1, \ldots, n$ are in the vaccine arm and individuals $i = n+1, \ldots, N$ are in the control arm. Henceforth, unless stated otherwise, time refers to the time since the start of the at-risk period, i.e. the time since the given individual reached 21 days after receipt of the second vaccine dose.

For individual $i = 1, \ldots, n$ in the vaccine arm, let $A_i(t)$ be the true antibody level at time $t$. Let $Y_i(t_{ij})$ be the observed antibody level at time $t_{ij}$, $j = 1, \ldots, m_i$. Note $m_i$, the number of antibody observations for individual $i$, may be 0. Assume observation $\log Y_i(t_{ij})$ is given by

$$\log Y_i(t_{ij}) = \log A_i(t_{ij}) + \epsilon_{ij} \tag{1}$$

where random error $\epsilon_{ij} \sim t_\nu(0, \sigma^2)$ follows the Student-t distribution with $\nu$ degrees of freedom, location 0 and scale $\sigma^2$. We used the Student-t distribution as QQ-plots showed better fit than using Gaussian error, and it is more robust to outliers than a Gaussian error distribution [1]. We chose $\nu = 4$ by examining QQ-plots of the residuals from an earlier model using Gaussian random error. We considered different integer values of $\nu$, and chose $\nu = 4$ as the QQ-plot was closest to a straight line.

Assume a linear trajectory of the true log antibodies $\log A_i(t)$ over time

$$\log A_i(t) = b_{0i} - \exp(b_{1i})\, t \tag{2}$$

We let $a_{0i} = b_{0i}$ and $a_{1i} = -\exp(b_{1i})$ be the random intercept and slope respectively. Random effects $b_{0i}$, $b_{1i}$ follow a Gaussian distribution given by

$$\begin{pmatrix} b_{0i} \\ b_{1i} \end{pmatrix} \sim N\left( \begin{pmatrix} \alpha_0 + x_{Li}^\top \beta_{L0} \\ \alpha_1 + x_{Li}^\top \beta_{L1} \end{pmatrix}, \begin{pmatrix} \tau_0^2 & \rho\tau_0\tau_1 \\ \rho\tau_0\tau_1 & \tau_1^2 \end{pmatrix} \right) \tag{3}$$

independently for each $i$. Here $X_L$ is a design matrix of $p_L$ covariates which affect the antibody trajectories with $i$th row $x_{Li}^\top$, and corresponding effects on the intercept and slope given by $\beta_{L0}$, $\beta_{L1}$ respectively. Parameters $\alpha_0$ and $\alpha_1$ give the expected random effects for a reference individual with $x_{Li} = 0$, $\tau_0^2$ and $\tau_1^2$ give the variances of the random effects, and $\rho$ the correlation between the random effects.

The use of the $-\exp$ transformation forces the log antibody slopes to be negative. We used this for two reasons: (a) a positive slope means antibody levels increase exponentially, which causes convergence issues in the time-to-event component of the model, (b) once antibody levels have reached their peak we expect them to decrease over time.

The formulation of equations (2) and (3) can be alternatively written as

$$\log A_i(t) = \alpha_0 + x_{Li}^\top \beta_{L0} + \tau_0 \eta_{0i} - t \exp\left(\alpha_1 + x_{Li}^\top \beta_{L1} + \tau_1 \left(\rho \eta_{0i} + \sqrt{1-\rho^2}\eta_{1i}\right)\right) \quad (4)$$

where $\eta_{0i}$, $\eta_{1i}$ are independent standard normal random variables, $i = 1, \ldots, n$. This formulation is used in the code implementing the model.

For individual $i = n+1, \ldots, N$ in the control arm, we assume their antibody levels are zero, $A_i(t) \equiv 0$.

##### 1.1.2 Time-to-infection component

We model the time until SARS-CoV-2 infection by the Cox proportional hazards model [2]. We consider the event of interest to be either any COVID-19 infection, or primary symptomatic COVID-19 infection, in two separate analyses. For individual $i$, let $S_i$ be the calendar time when they reached 21 days after their second dose, $T_i$ the total time at risk, and $\delta_i = \left(\delta_i^{(\text{any})}, \delta_i^{(\text{symp})}\right)^\top$ the vector of event indicators for any COVID-19 infection and symptomatic COVID-19 infection respectively. Let $Z_i = 1$ if individual $i$ is in the vaccine arm, and $Z_i = 0$ if they are in the control arm. Let $X_I$ be a design matrix of covariates which affect the risk of infection, with row $x_{Ii}^\top$ for the $i$th individual. We stratify the baseline hazard function by study site $G_i$, as the rate of COVID-19 in the population varied by geographical study site during the trial. We assume a 7 day incubator period from exposure to a COVID-19 infection being reported in the study. Hence the risk of infection at a given time depends on the true antibody levels 7 days prior. Then the hazard $h_i(t)$ for individual $i$ at time $t$ depends on the vaccine/control arm $Z_i$, the true antibody level $A_i(t-7)$, and covariates $x_{Ii}$

$$h_i(t) = \lambda_{G_i}(S_i + t) \exp\left(x_{Ii}^\top \beta_I + Z_i\left(\gamma A_i(t-7) + \zeta\right)\right) \quad (5)$$

where $G_i$ is the study site for individual $i$, $\lambda_{G_i}(s)$ is the unknown baseline hazard function for study site $G_i$ which varies with calendar time, $\beta_I$ is the effect of covariates $x_{Ii}$ on the log hazard, $\gamma$ is the slope of the antibody level effect $A_i(t-7)$, and $\zeta$ is the intercept of the antibody level effect.

Note the antibody levels $A_i(t)$ decay to 0 as $t \to \infty$. Hence the hazard ratio between an individual in the vaccine arm and control arm with the same covariate values will tend to $\exp(\zeta)$ as $t \to \infty$, so we can interpret $\exp(\zeta)$ as the long-term effect due to vaccination.

We implement the longitudinal component of the model using Bayesian methods. This allows us to easily deal with the missing antibody data by treating the antibody trajectory parameters as latent variables. We then multiply impute the latent true antibody trajectories, before using a Cox model to implement the time-to-infection component.

#### 1.2 Bayesian implementation of longitudinal component

##### 1.2.1 Covariates

In equation (3), we include covariates in $X_L$ which may affect the initial antibody level and slope of antibody decay after vaccination. We include categorical variables: age group (18-55 years, 56-69 years, 70+ years), sex (female, male), ethnicity (white, non-white), comorbidity (none, comorbidity), BMI (<30 kg/m$^2$, $\geq$30), time interval between first and second dose ($\geq$12 weeks, 9-11 weeks, 6-8 weeks, <6 weeks), healthcare worker (HCW) status (not a HCW, HCW facing <1 COVID-19 patient per day, HCW facing $\geq$1 COVID-19 patient per day), and initial dose (standard dose, low dose).

##### 1.2.2 Accounting for informative missingness

Missing data occurred in the antibody measurements for two reasons. Firstly, longitudinal antibody observations were censored, that is, excluded if they occur after the infection event or the end of the at-risk period. Secondly, blood samples were tested for antibody levels retrospectively by case-cohort sampling, meaning antibody data is missing for a large proportion of non-cases. Hence the missingness of antibody data depends by design on the total time at risk and the final infection status. Because antibody levels affect the risk of infection, these time-to-event variables are informative of the antibody levels. In order to account for this informatively missing-at-random antibody data, we also included infection data in the longitudinal covariates. Namely, we included the indicator for any COVID-19 infection, the indicator for primary symptomatic COVID-19 infection, and the Nelson–Aalen estimate for cumulative hazard of any COVID-19 NAAT+ test in the vaccine group. This approach was first suggested for the Cox model by White and Royston (2009) [3]. Moreno-Bentacur et al. (2018) used a similar approach in a joint model, suggesting to also include the interaction between other covariates and the Nelson–Aalen estimator, although they note the interaction term did not improve the estimates [4]. The inclusion of both infection indicators accounts for the censoring at any infection event. This can further be seen as a multiple imputation approach to fitting a joint model with two competing risks of non-symptomatic COVID-19 infection and symptomatic COVID-19 infection.

##### 1.2.3 Covariate effects

The multiplicative effect due to an increase of 1 in the $j$th covariate $x_{Lj}$ on the antibody level at PB28 and the half-life of antibody level decay is given by $\exp(\beta_{L0j})$ and $\exp(-\beta_{L1j})$ respectively. This can be derived from Equation (4).

##### 1.2.4 Standardisation

We standardised the data before estimating the posterior, in order to improve mixing in the Hamiltonian Monte Carlo algorithm. We recentered the time since second vaccination variable at 28 days post second dose, and divided by the standard deviation of the antibody observation times. We standardised the covariates and observed antibody levels by subtracting the mean and dividing by the standard deviation of each variable. We then fit the model on the standardised data, before transforming the parameters back to the original scale.

###### 1.2.5 Priors

We fit the following independent priors to the parameters, where $\theta'$ indicates the parameter $\theta$ transformed to the standardised scale:

$$\alpha_0',\ \alpha_1' \sim \text{Normal}(0, 2^2) \tag{6}$$

$$\beta_{L0}', \beta_{L1}' \sim \text{Normal}(0, 2^2 I_{p_L}) \tag{7}$$

$$\tau_0',\ \tau_1',\ \sigma' \sim \text{Student-t}(\text{d.f.} = 2, \text{mean} = 0, \text{scale} = 1) \tag{8}$$

$$\rho' \sim \text{Uniform}(-1, 1) \tag{9}$$

where $I_{p_L}$ denotes the identity matrix with $p_L$ rows. These priors, combined with equations (2)-(3) induce a prior on $A_i(t)$.

###### 1.2.6 Model fitting

We implement the model using Hamiltonian Monte Carlo [5] in Stan [6]. We ran 4 chains of 20000 iterations each, discarding the first 5000 iterations as warm-up and combining all the remaining iterations from each chain. We checked for convergence by examining trace plots and checking all values of the potential scale reduction factor were less than 1.01.

##### 1.3 Approximate Bayesian inference procedure

We estimate the joint posterior distribution by multiple imputation.

The joint posterior distribution for the antibody trajectories $\boldsymbol{A}(t) = (A_1(t), \ldots, A_n(t))^\top$ and time-to-event parameters $\theta_I = (\gamma, \zeta, \beta_I)^\top$ is given by

$$\pi\left(\boldsymbol{A}(t), \theta_I \,|\, Y, T, \delta, X_L, X_I\right) = \pi\left(\theta_I \,|\, \boldsymbol{A}(t), Y, T, \delta, X_L, X_I\right)\ \pi\left(\boldsymbol{A}(t) \,|\, Y, T, \delta, X_L, X_I\right) \tag{10}$$

Hence to sample from the joint posterior distribution, we may first draw a sample of antibody trajectories $\boldsymbol{A}(t)^{(k)} \sim \pi\left(\boldsymbol{A}(t) \,|\, Y, T, \delta, X_L, X_I\right)$, then draw a sample of time-to-event parameters $\theta_I^{(k)} \sim \pi\left(\theta_I \,|\, \boldsymbol{A}(t), Y, T, \delta, X_L, X_I\right)$, given the sample of antibody trajectories. We approximate the distribution of antibody trajectories given the antibody and time-to-event data $\pi\left(\boldsymbol{A}(t) \,|\, Y, T, \delta, X_L, X_I\right)$ in Section 1.2. Note Section 1.2.2 means our samples are drawn from an approximation of the appropriate posterior, which conditions on the time-to-event data $T, \delta$.

We first impute the latent true antibody trajectories from the Bayesian posterior defined in Section 1.2. We then run the time-to-event model to estimate the distribution of the time-to-event parameters given the imputation.

For $k = 1, \ldots, K$, we impute the latent values of the true antibody level using draws of the random intercept $a_{0i}^{(k)}$ and random slope $a_{1i}^{(k)}$, $i = 1, \ldots, n$ from the posterior of the longitudinal model. A set of imputed intercepts and slopes $a_0^{(k)}$, $a_1^{(k)}$ defines the true antibody trajectories $A_i^{(k)}(t),\ i = 1, \ldots, n$ in Eq. (5).

We then run a Cox proportional hazards model with time-varying covariate $A_i^{(k)}(t-7)$ and hazard given by Eq. (5). The model gives us the maximum likelihood estimate (MLE) $\hat{\theta}_I^{(k)} = \left(\hat{\gamma}^{(k)}, \hat{\zeta}^{(k)}\right)^\top$

and corresponding covariance matrix $\widehat{\text{Var}}^{(k)}_{\theta_I}$ for the time-to-infection parameters, given the $k$th imputation of antibody trajectories.

Next we sample from the asymptotic distribution of the Cox maximum likelihood estimators $\hat{\theta}^{(k)}_I$, such that $\theta^{(k)}_I \sim N\left(\hat{\theta}^{(k)}_I, \widehat{\text{Var}}^{(k)}_{\theta_I}\right)$. This approximates a Bayesian posterior with diffuse improper priors [7], namely the distribution of $\pi\left(\theta^{(k)}_I \,|\, \boldsymbol{A}(t)^{(k)}, Y, T, \delta, X_L, X_I\right)$ in Eq. (10). Then our samples $\theta^{(k)}_I$, $\boldsymbol{A}^{(k)}(t)$ are approximate draws from the joint posterior $\pi(\theta_I, \boldsymbol{A}(t) \,|\, Y, T, \delta, X_L, X_I)$. Posterior quantities of interest can be calculated from these samples and summarised by posterior medians, means, and 95% credible intervals by calculating quantiles.

#### 1.4 Calculation of posterior quantities

##### 1.4.1 Vaccine efficacy at a given antibody level

The vaccine efficacy at a given antibody level $A$ is given by

$$\text{VE(A)} = 1 - \exp(\gamma \text{A} + \zeta) \tag{11}$$

##### 1.4.2 Antibody corresponding to a given vaccine efficacy

The antibody level $A$ required for a given vaccine efficacy VE is then given by the inverse of Eq. (11)

$$\text{A(VE)} = \left(\log(1 - \text{VE}) - \zeta\right)/\gamma \tag{12}$$

When this is negative, we report antibody of 0, as negative antibodies are not possible.

##### 1.4.3 Mean vaccine efficacy over time

The mean vaccine efficacy at time $t$ is then given by

$$\text{VE}(t) = \frac{1}{n}\sum_{i=1}^{n} \text{VE}(\text{A}_i(t-7)) = 1 - \frac{1}{n}\sum_{i=1}^{n} \exp(\gamma \text{A}_i(t-7) + \zeta) \tag{13}$$

Note this is describing a counterfactual scenario where individuals are exposed to COVID-19 at time $t$ and not exposed prior. In real-life populations, individuals who test positive are protected from future infection (for a period) and drop out from the sum. This will be more likely for individuals who are higher risk for COVID-19, including namely unvaccinated individuals. Hence the observed vaccine efficacy in a population will be shrunk towards a null effect compared to the counterfactual vaccine efficacy we report here. This is sometimes known as the differential depletion of susceptibles bias [8].

##### 1.4.4 Quantiles of vaccine efficacy

Let $\mathbf{A}(t)$ be the vector with $i$th entry $\mathrm{A}_i(t)$. Let $Q(\boldsymbol{x}, q)$ be the $q$th quantile of the vector $\boldsymbol{x}$. Then the $q$th quantile of vaccine efficacy at time $t$, $\mathrm{VE}_q$ is then given by

$$\mathrm{VE}_q(t) = Q\left(\mathrm{VE}(\mathbf{A}(t-7)), q\right) = 1 - \exp(\gamma Q(\mathbf{A}(t-7), q) + \zeta) = \mathrm{VE}(Q(\mathbf{A}(t-7), q)) \qquad (14)$$

That is, the $q$th quantile of vaccine efficacy at time $t$ is equal to the vaccine efficacy given at the $q$th quantile of antibody levels at time $t-7$. As in Section 1.4.3 above, this calculates a counterfactual vaccine efficacy, in which individuals are not exposed to the virus until time $t$. In an observed population individuals who have the lowest antibody response will be infected earlier on average, and drop out of the at-risk group.

##### 1.4.5 Calculating covariate effects on vaccine efficacy

To calculate covariate effects on vaccine efficacy, we predicted efficacy in new individuals with given covariate values. As mentioned in the main text, the event indicators and Nelson–Aalen (N–A) estimate of cumulative hazard for a new individual are unknown, as they depend on the infection outcome. We first built an imputation model to sequentially impute the missing event indicators (infection, symptomatic) and N–A estimate, which appear in the longitudinal antibody model, given the known covariates.

**Imputing the missing time-to-event data** We imputed the missing time-to-event data sequentially. We ran regression models on the observed dataset, then drew predictions from them for the new individuals to impute the missing values.

We first ran a logistic model to predict the probability of any COVID-19 infection using all longitudinal covariates $x_L$ except the event indicators and Nelson–Aalen estimate (logmI). We then ran a logistic model to predict the probability of primary symptoms given infection occurred, using all longitudinal covariates $x_L$ except N–A estimate (logmS). We then ran a linear model to predict the N–A estimate using all longitudinal covariates $x_L$ including infection and primary symptomatic status (linmNA).

For a new individual, we imputed the infection indicator to be 1 with probability equal to their predicted probability of infection from the logmI model above, and 0 otherwise. If the imputed infection indicator is 1, we next imputed the symptomatic infection indicator to be 1 with probability equal to their predicted probability of symptomatic infection given they were infected, predicted from the logmS model above, and 0 otherwise. Given the imputed event indicators, we imputed the N–A estimate to be the predicted value from the linear model linmNA, plus a randomly sampled residual from linmNA. If the resulting imputated value is less than 0, we replace it with 0, as the N–A estimate is non-negative by definition. Together, these imputations are an approximate draw from the distribution of the event indicators and N–A estimate given the longitudinal covariates.

**Predicted vaccine efficacy for a new individual** We imputed these missing covariates $n$ times. For each set of imputed covariates, we drew from the predictive distribution of antibody trajectories for a new individual with the given covariate values, $A_j^{\text{pred}}(t), j = 1, \ldots, n$. We then averaged these to calculate the mean risk of infection given the known covariate values

$$\text{VE}(t) = \frac{1}{n}\sum_{j=1}^{n} \text{VE}(A_j^{\text{pred}}(t)) = 1 - \frac{1}{n}\sum_{j=1}^{n} \exp(\gamma A_j^{\text{pred}}(t) + \zeta) \tag{15}$$

##### 1.4.6 Predicted relative vaccine efficacy against Omicron BA.4/5 infection

To estimate the relative efficacy against Omicron BA.4/5 infection over time compared with vaccine non-responders, we considered three scenarios. We assumed the estimated mean, 95% upper and lower CI bounds for the relationship between anti-spike IgG antibody levels and relative efficacy against Omicron BA.4/5 infection from Wei et al. (2023) [9] to be the true relationship in each scenario respectively. We extracted the estimate and 95% CI from Wei et al. (Supplementary Fig. 1 from Wei et al.), at every 500 BAU/mL (0, 500, 1000, 1500, . . . , 8000), and at every 10% relative efficacy (0%, 10%, 20%, . . . , 80%). We then fitted a LOESS curve to each of the three curves (95% lower confidence bound, estimate, 95% upper confidence bound), with a span of 0.3. The span was chosen such that the curve visually fit the data well, with no signs of overfitting. We then predicted the relative efficacy against Omicron infection RE(A) at a given antibody level A from the fitted LOESS curve. The mean relative efficacy against Omicron in the trial is then given by

$$\text{RE}(t) = \frac{1}{n}\sum_{i=1}^{n} \text{RE}(A_i(t-7)) \tag{16}$$

### 2 Supplementary Results

#### 2.1 Predicted efficacy over time against the Omicron BA.4/5 variant

We extrapolated our results to predict what the observed relative efficacy over time in our study might have been, were the trial conducted during the Omicron period. We predicted efficacy relative to vaccine non-responders, defined below. Wei et al. (2023) estimated the relationship between anti-spike IgG antibody levels and relative efficacy against Omicron BA.4/5 variant infection after booster vaccination following two initial doses of COVID-19 vaccine [9]. They calculated efficacy relative to vaccinated individuals who produced antibody levels of 16 BAU/mL, deemed a non-response. We applied their estimated relationship between anti-spike IgG antibody levels and relative efficacy against the Omicron BA.4/5 variant to the predicted antibody levels over time from this study. We considered three scenarios: assuming their reported mean, upper- and lower 95% confidence limit for the relationship between anti-spike IgG and relative efficacy against Omicron BA.4/5 to be the true relationship. We refer to these as the mean efficacy, high efficacy and low efficacy scenarios respectively. This analysis assumes the relationship between antibody levels and efficacy after two doses of ChAdOx1 nCoV-19 is the same as the relationship after three vaccine doses (of possibly different vaccines). This assumption is unlikely to hold, which may bias our analysis (see Supplementary Discussion). In the mean efficacy scenario, we estimated relative efficacy at day 35, 97 and 189 to be 18.5% (95% CrI: 17.8, 19.2), 11.0% (10.5, 11.6) and 4.9% (4.4, 5.5) respectively (Supplementary

Fig. 11). In the high and low efficacy scenarios respectively, we estimate relative efficacy of 23.4% (22.6, 24.2) and 13.6% (12.9, 14.2) at day 35, 14.8% (14.1, 15.4) and 7.4% (7.0, 7.9) at day 97 and 7.1% (6.5, 7.8) and 3.1% (2.7, 3.5) at day 189. The credible intervals do not reflect the uncertainty in the relationship between antibody levels and relative efficacy. This indicates the antibodies induced by two doses of ChAdOx1 nCoV-19 might only induce minimal protection against Omicron BA.4/5 of at most 24.2% at day 35, waning to at most 7.8% at day 189, relative to a vaccinated individual with no antibodies (see Discussion).

#### 2.2 Model fit

To demonstrate how the model works, Supplementary Fig. 12 shows the estimated anti-spike IgG antibody trajectories over time for 12 different randomly chosen individuals We randomly chose 12 individuals, 3 each with 0, 1, 2 and 3 antibody observations available respectively, and plotted their estimated true anti-spike IgG levels over time. The plot demonstrates that individuals with no antibody observations are predicted to have a typical antibody trajectory, with a large uncertainty. Individuals with more observations have less uncertainty around their estimated antibody levels over time.

#### 2.3 Computation time

The longitudinal model was run in Stan with 4 chains, in parallel on a SLURM compute cluster. This took 2h 23m (9h 33m total walltime), and required 52GB of RAM to generate 15000 iterations per chain, after 5000 iterations of warmup. The Cox model was run in parallel using the snowfall package in R, on the imputed datasets from each iteration from the longitudinal model. This was run on a SLURM compute cluster separately for each outcome (symptomatic, all infections). Each outcome was run using 24 parallel nodes, taking 2h 37m (2 days 12h, 6m total walltime) and 2h 43m (2 days 12h, 35m total walltime) for the symptomatic and all infection analyses respectively. The Cox analysis required 41GB and 38GB of RAM respectively. The results from the Cox and longitudinal models were combined to obtain the vaccine efficacy results reported in the paper in a third step using R and the snowfall package. This was run on 24 parallel nodes on a SLURM compute cluster, taking 1h 28m (1 day 11h 10m total walltime), and 1h 22m (1 day 8h 50m total walltime) for the symptomatic and all infection outcomes respectively. The final analysis required 49GB and 50GB of RAM respectively.

#### 2.4 Calculated parameters

Posterior summaries of the estimated parameters are reported as follows. The intercept, slope and related variance and correlation parameters are reported in Supplementary Table 4. The multiplicative covariate effects on 28 days post second dose and half-life are reported in Fig. 4 and Supplementary Table 5. These are given by $\exp(\beta_0)$ and $\exp(-\beta_1)$ respectively, and the natural logarithm can be applied to calculate $\beta_0$ and $\beta_1$. For the time-to-event model, the covariate effects on hazard (including antibody and vaccination) are reported in Supplementary Fig. 7, and Supplementary Table 6.

### 3 Supplementary Discussion

Our data was collected in a period when the Alpha variant was dominant, and considered individuals without prior infection who received two doses of vaccine. In contrast, the Omicron variant is currently dominant, and many individuals have received three or more doses of vaccine and been previously infected. While some findings such as effects of dose interval on vaccine efficacy will likely still apply, our results are therefore not directly applicable to current and future populations. To aid with this, we extrapolated our results to examine what the efficacy over time since second dose might have been in the Omicron BA.4/5 period. We used the estimated relationship between efficacy and antibody levels reported by Wei et al. (2023) [9] to estimate the level of Omicron BA.4/5 protection induced by antibody trajectories observed in this trial. This analysis should be interpreted with caution for three reasons: firstly, Wei et al., and thus also our analysis, use vaccine non-responders (vaccinated individuals whose antibody level was 16 BAU/mL) as the comparison in their calculation of relative efficacy instead of unvaccinated controls. It is not clear therefore how our analysis, or that of Wei et al., might relate to protection against Omicron BA.4/5 compared with unvaccinated individuals. Secondly, the relationship reported by Wei et al. was calculated from observational data, which may introduce bias into our analysis. Thirdly, our analysis assumed that the relationship between antibody levels and efficacy was the same after two doses in our study, as in the Wei et al. study. Their study combined individuals vaccinated with two, three or four vaccine doses (83.9% had three doses), including individuals who received the ChAdOx1 nCoV-19 and BNT162b2 as their first two doses, and different booster dose vaccines. The relationship between antibody levels and efficacy may differ across these groups, which may introduce some bias into our analysis. We suggest that protection from two doses of vaccine is likely to be no greater than after three doses, at the same level of antibodies. Therefore any such bias is likely to result in our reported estimates being an over-estimate of the effectiveness of two doses against Omicron BA.4/5, compared with vaccine non-responders. These estimates are all calculated for individuals with no prior infection, which is rare today. Levels of vaccine-induced protection may be different for individuals with a prior infection.

We estimated low efficacy after two doses of ChAdOx1 nCoV-19 vaccine against the Omicron BA.4/5 variant, of 18.5% at 35 days, 7.6% at 140 days and 4.1% at 209 days, relative to vaccine non-responders (Supplementary Fig. 11). Supplementary Table 8 compares our methods and results to previous studies of vaccine efficacy against the Omicron variant. Andrews et al. (2022b) used a test-negative design to estimate VE against Omicron [10] and is vulnerable to the aforementioned biases from observational studies. Andrews et al. reported a much higher vaccine efficacy against Omicron at 2-4 weeks since second dose, but report a small negative vaccine efficacy at 25 weeks since second dose, which may be due to bias. While the results from our study are not directly comparable with theirs, it is however interesting that they reported higher initial VE against Omicron than our estimated relative efficacy, with both studies waning to negligible protection after 25 weeks. This might indicate that vaccine non-responders are initially protected against infection relative to vaccine non-responders, but this protection wanes by 25 weeks.

### 4 Supplementary Tables

**Supplementary Table 1: Baseline characteristics of vaccinated (ChAdOx1) and control arms**

| | ChAdOx1 (n=4605) | Control (n=4423) |
|---|---|---|
| Age (years) | | |
| Median (IQR) | 45 (34, 56) | 45 (34, 55) |
| 18-55 years | 3435 (74.6%) | 3423 (77.4%) |
| 55-69 years | 568 (12.3%) | 503 (11.4%) |
| 70+ years | 602 (13.1%) | 497 (11.2%) |
| Sex | | |
| Male | 1934 (42.0%) | 1765 (39.9%) |
| Female | 2671 (58.0%) | 2658 (60.1%) |
| Ethnicity | | |
| White | 4235 (92.0%) | 4092 (92.5%) |
| Other | 370 (8.0%) | 331 (7.5%) |
| Comorbidities | | |
| None | 3475 (75.5%) | 3348 (75.7%) |
| Comorbidity | 1130 (24.5%) | 1075 (24.3%) |
| BMI ($kg/m^2$) | | |
| Median (IQR) | 25.5 (23, 28.8) | 25.4 (22.9, 28.9) |
| BMI $<30$ $kg/m^2$ | 3704 (80.4%) | 3531 (79.8%) |
| BMI $\geq30$ $kg/m^2$ | 901 (19.6%) | 892 (20.2%) |
| Healthcare worker status | | |
| Non-healthcare worker | 1759 (38.2%) | 1550 (35.0%) |
| Healthcare worker (<1 patient with COVID-19 per day) | 1973 (42.8%) | 2008 (45.4%) |
| Healthcare worker (≥1 patient with COVID-19 per day) | 873 (19.0%) | 865 (19.6%) |
| Interval between first and second dose | | |
| Median (IQR) days | 74 (42, 89) | 76 (49, 89) |
| <6 weeks | 1722 (37.4%) | 1673 (37.8%) |
| 6-8 weeks | 1215 (26.4%) | 1294 (29.3%) |
| 9-11 weeks | 555 (12.1%) | 497 (11.2%) |
| ≥12 weeks | 1113 (24.2%) | 959 (21.7%) |
| First dose strength | | |
| Standard Dose | 2979 (64.7%) | 2913 (65.9%) |
| Low Dose | 1626 (35.3%) | 1510 (34.1%) |
| NAAT+ case* | | |
| Negative | 4398 (95.5%) | 4046 (91.5%) |
| NAAT+ case | 207 (4.5%) | 377 (8.5%) |
| Primary symptomatic NAAT+ case† | | |
| Non-case | 4534 (98.5%) | 4206 (95.1%) |
| NAAT+ primary symptomatic case | 71 (1.5%) | 217 (4.9%) |
| Time from day 28 post second dose until infection | | |
| Median (IQR) days | 64 (38.5, 98) | 59 (35, 91) |
| Total follow up time at risk | | |
| Median (IQR) days | 102 (77, 128) | 97 (73, 122) |

Values are counts (percentages) unless stated otherwise. *NAAT+ case denotes nucleic acid amplification test positive participants. †Primary symptomatic NAAT+ denotes nucleic acid amplification test positive participants with at least one qualifying symptom.

**Supplementary Table 2: Observed anti-spike IgG antibody measurements by visit, arm (ChAdOx1/Control) and case**

| | | ChAdOx1 | | | | Control |
|---|---|---|---|---|---|---|
| | | Non-case<br>n=4398 (95.5%) | Case<br>n=207 (4.5%) | Symptomatic case<br>n=71 (1.5%) | Total<br>n=4605 | Total<br>n=4423 |
| PB28 | Number of antibody measurements available | 1155 (26.3%) | 173 (83.6%) | 59 (83.1%) | 1328 (28.8%) | 852 (19.3%) |
| | Median (IQR) anti-spike IgG value BAU/mL | 219 (119, 383) | 197 (103, 322) | 165 (100, 276) | 214 (115, 370) | 0.3 (<LLOQ, 0.5) |
| | Number of antibody measurements <LLOQ | 0 (0%) | 0 (0%) | 0 (0%) | 0 (0%) | 357 (41.9%) |
| PB90 | Number of antibody measurements available | 519 (11.8%) | 64 (30.9%) | 23 (32.4%) | 583 (12.7%) | 62 (1.4%) |
| | Median (IQR) anti-spike IgG value BAU/mL | 115 (66, 206) | 93 (47, 168) | 78 (62, 178) | 112 (64, 199) | 0.3 (<LLOQ, 0.6) |
| | Number of antibody measurements <LLOQ | 0 (0%) | 0 (0%) | 0 (0%) | 0 (0%) | 21 (33.9%) |
| PB182 | Number of antibody measurements available | 58 (1.3%) | 1 (0.5%) | 1 (1.4%) | 59 (1.3%) | 9 (0.2%) |
| | Median (IQR) anti-spike IgG value BAU/mL | 65 (45, 117) | 30 (NA) | 30 (NA) | 64 (44, 117) | 0.2 (<LLOQ, 0.3) |
| | Number of antibody measurements <LLOQ | 0 (0%) | 0 (0%) | 0 (0%) | 0 (0%) | 4 (44.4%) |
| Total | Number of antibody measurements available | 1732 (39.4%) | 238 (115%) | 83 (116.9%) | 1970 (42.8%) | 923 (20.9%) |
| | Median (IQR) anti-spike IgG value BAU/mL | 175 (92, 320) | 164 (82, 280) | 137 (69, 243) | 172 (91, 312) | 0.3 (<LLOQ, 0.5) |
| | Number of antibody measurements <LLOQ | 0 (0%) | 0 (0%) | 0 (0%) | 0 (0%) | 382 (41.4%) |

Only ChAdOx1 vaccinated participants are split by case (Non-case, Case, Symptomatic case). The number of antibody measurements available, median and interquartile range, and number of measurements below the lower limit of quantification (LLOQ) 0.21 BAU/mL is shown. Antibody measurements are given in BAU/mL to the nearest whole number for the ChAdOx1 arm, and to 1 decimal place for the Control arm. Outliers are not included in this table (see Supplementary Table 3).

**Supplementary Table 3: The 8 outlying antibody values from ChAdOx1 vaccinated participants which were removed from the analysis**

| Case | Days since second dose | Visit | Anti-spike IgG value (BAU/mL) |
|---|---|---|---|
| Non-case | 40 | PB28 | <LLOQ |
| Non-case | 34 | PB28 | 0.6 |
| Non-case | 27 | PB28 | 0.3 |
| Non-case | 89 | PB90 | <LLOQ |
| Non-case | 103 | PB90 | 4363.8 |
| Symptomatic case | 95 | PB90 | 3.3 |
| Non-case | 91 | PB90 | 0.7 |
| Non-case | 91 | PB90 | <LLOQ |

Values less than the lower limit of quantification 0.21 BAU/mL are written as $<$ LLOQ. All except the 5th value were removed due to being extremely low, which may lead to issues in the model fit, encouraging the variance of random intercepts and slopes to be too large (See Supplementary Fig. 2)

**Supplementary Table 4: Posterior summary of parameters in the longitudinal antibody model, for antibody measurements converted to BAU/mL**

| Parameter | Mean | Median | 0.025 quantile | 0.975 quantile |
|---:|---:|---:|---:|---:|
| $\alpha_0$ | 5.761 | 5.762 | 5.513 | 6.006 |
| $\alpha_1$ | $-4.770$ | $-4.770$ | $-5.032$ | $-4.506$ |
| $\sigma_e$ | 0.206 | 0.206 | 0.184 | 0.226 |
| $\tau_0$ | 0.756 | 0.756 | 0.722 | 0.791 |
| $\tau_1$ | 0.162 | 0.170 | 0.014 | 0.287 |
| $\rho$ | $-0.134$ | $-0.103$ | $-0.767$ | 0.269 |

Time $t = 0$ is set to be 28 days after the second dose in the model implementation. A reference individual in the model is one with age 18-55, male, white, no comorbidity, $\geq$ 12 weeks interval between first and second dose, BMI $< 30\text{kg/m}^2$, not a healthcare worker, standard first dose, negative COVID-19 and 0 Nelson–Aalen estimate.

**Supplementary Table 5: The multiplicative covariate effects on anti-spike IgG level 28 days post second dose, and half-life of the subsequent decay in anti-spike IgG levels**

| Covariate multiplicative effect | Posterior estimates | | | |
|---|---|---|---|---|
| Multiplicative effect on anti-spike IgG level 28 days post second dose | Median | Mean | 2.5% quantile | 97.5% quantile |
| Age (56-69 vs 18-55 years) | 0.86 | 0.86 | 0.71 | 1.03 |
| Age ($\geq$70 vs 18-55 years) | 0.78 | 0.78 | 0.63 | 0.96 |
| Sex (Female vs Male) | 1.08 | 1.08 | 0.99 | 1.19 |
| Ethnicity (Non-white vs White) | 1.18 | 1.18 | 1.00 | 1.39 |
| Comorbidity (Comorbidity vs None) | 1.00 | 1.00 | 0.90 | 1.11 |
| BMI ($\geq$30 vs $<$30 kg/m$^2$) | 1.00 | 1.00 | 0.90 | 1.12 |
| Healthcare worker (HCW) status (HCW facing $<$1 COVID-19 patient per day vs Not a HCW) | 0.84 | 0.84 | 0.73 | 0.95 |
| Healthcare worker (HCW) status (HCW worker facing $\geq$1 COVID-19 patient per day vs Not a HCW) | 0.90 | 0.90 | 0.77 | 1.06 |
| Interval between first and second dose (9-11 weeks vs $\geq$12 weeks) | 0.83 | 0.83 | 0.74 | 0.93 |
| Interval between first and second dose (6-8 weeks vs $\geq$12 weeks) | 0.70 | 0.70 | 0.61 | 0.81 |
| Interval between first and second dose ($<$6 weeks vs $\geq$12 weeks) | 0.51 | 0.52 | 0.43 | 0.61 |
| First dose (Low dose vs Standard dose) | 1.11 | 1.11 | 0.99 | 1.24 |
| COVID-19 outcome (Positive vs Negative) | 0.85 | 0.86 | 0.71 | 1.01 |
| COVID-19 outcome (Primary symptomatic vs Negative) | 0.76 | 0.76 | 0.61 | 0.95 |
| Cumulative hazard (Nelson–Aalen estimator for cumulative hazard) | 0.97 | 0.97 | 0.92 | 1.03 |
| Multiplicative effect on anti-spike IgG level half-life | Median | Mean | 2.5% quantile | 97.5% quantile |
| Age (56-69 vs 18-55 years) | 1.22 | 1.25 | 0.89 | 1.74 |
| Age ($\geq$70 vs 18-55 years) | 0.86 | 0.86 | 0.62 | 1.15 |
| Sex (Female vs Male) | 0.90 | 0.90 | 0.81 | 0.99 |
| Ethnicity (Non-white vs White) | 1.09 | 1.09 | 0.90 | 1.34 |
| Comorbidity (Comorbidity vs None) | 0.92 | 0.92 | 0.82 | 1.04 |
| BMI ($\geq$30 vs $<$30 kg/m$^2$) | 0.84 | 0.84 | 0.74 | 0.94 |
| Healthcare worker (HCW) status (HCW facing $<$1 COVID-19 patient per day vs Not a HCW) | 0.96 | 0.96 | 0.83 | 1.11 |
| Healthcare worker (HCW) status (HCW worker facing $\geq$1 COVID-19 patient per day vs Not a HCW) | 0.88 | 0.88 | 0.76 | 1.03 |
| Interval between first and second dose (9-11 weeks vs $\geq$12 weeks) | 0.99 | 0.99 | 0.88 | 1.11 |
| Interval between first and second dose (6-8 weeks vs $\geq$12 weeks) | 0.97 | 0.97 | 0.84 | 1.12 |
| Interval between first and second dose ($<$6 weeks vs $\geq$12 weeks) | 0.96 | 0.98 | 0.75 | 1.27 |
| First dose (Low dose vs Standard dose) | 1.09 | 1.09 | 0.97 | 1.23 |
| COVID-19 outcome (Positive vs Negative) | 0.86 | 0.86 | 0.72 | 1.04 |
| COVID-19 outcome (Primary symptomatic vs Negative) | 0.78 | 0.78 | 0.64 | 0.97 |
| Cumulative hazard (Nelson–Aalen estimator for cumulative hazard) | 1.01 | 1.01 | 0.96 | 1.07 |

A forest plot is given in Fig. 4

**Supplementary Table 6: The multiplicative covariate effects on hazard against symptomatic COVID-19 infection and any COVID-19 infection**

| Covariate multiplicative effect | Posterior estimates | | | |
|---|---|---|---|---|
| Multiplicative effect on hazard for symptomatic COVID-19 infection | Median | Mean | 2.5% quantile | 97.5% quantile |
| Antibody level (Effect due to increase of 100 BAU/mL) | 0.34 | 0.32 | 0.08 | 0.76 |
| Vaccination direct effect (ChAdOx1 nCoV-19 vs control) | 0.71 | 0.73 | 0.38 | 1.53 |
| Age (56-69 vs 18-55 years) | 0.50 | 0.50 | 0.29 | 0.89 |
| Age (≥70 vs 18-55 years) | 0.16 | 0.16 | 0.06 | 0.42 |
| Sex (Female vs Male) | 0.88 | 0.88 | 0.68 | 1.13 |
| Ethnicity (Non-white vs White) | 1.02 | 1.02 | 0.67 | 1.56 |
| Comorbidity (Comorbidity vs None) | 0.98 | 0.98 | 0.73 | 1.32 |
| BMI (≥30 vs <30) | 1.39 | 1.39 | 1.06 | 1.82 |
| Healthcare worker (HCW) status (HCW facing <1 COVID-19 patient per day vs Not a HCW) | 1.42 | 1.42 | 1.00 | 2.02 |
| Healthcare worker (HCW) status (HCW worker facing ≥1 COVID-19 patient per day vs Not a HCW) | 2.20 | 2.20 | 1.49 | 3.24 |
| Multiplicative effect on hazard for any COVID-19 infection | Median | Mean | 2.5% quantile | 97.5% quantile |
| Antibody level (Effect due to increase of 100 BAU/mL) | 0.49 | 0.48 | 0.27 | 0.75 |
| Vaccination direct effect (ChAdOx1 nCoV-19 vs control) | 0.92 | 0.93 | 0.64 | 1.39 |
| Age (56-69 vs 18-55 years) | 0.66 | 0.66 | 0.46 | 0.93 |
| Age (≥70 vs 18-55 years) | 0.53 | 0.53 | 0.36 | 0.80 |
| Sex (Female vs Male) | 0.93 | 0.93 | 0.78 | 1.11 |
| Ethnicity (Non-white vs White) | 0.86 | 0.86 | 0.62 | 1.19 |
| Comorbidity (Comorbidity vs None) | 0.95 | 0.95 | 0.77 | 1.16 |
| BMI (≥30 vs <30) | 1.13 | 1.13 | 0.92 | 1.38 |
| Healthcare worker (HCW) status (HCW facing <1 COVID-19 patient per day vs Not a HCW) | 1.18 | 1.18 | 0.93 | 1.50 |
| Healthcare worker (HCW) status (HCW worker facing ≥1 COVID-19 patient per day vs Not a HCW) | 1.65 | 1.65 | 1.26 | 2.17 |

A forest plot is given in Supplementary Fig. 7

**Supplementary Table 7: Anti-spike IgG antibody levels required to give different levels of vaccine efficacy against any COVID-19 infection or symptomatic COVID-19**

| | Required anti-spike IgG antibody level (BAU/mL) | |
|---|---|---|
| Vaccine efficacy (%) | Symptomatic COVID-19 | Any COVID-19 infection |
| 0 | 0 (0, 18) | 0 (0, 27) |
| 5 | 0 (0, 20) | 0 (0, 31) |
| 10 | 0 (0, 23) | 3 (0, 36) |
| 15 | 0 (0, 26) | 11 (0, 41) |
| 20 | 0 (0, 28) | 20 (0, 47) |
| 25 | 0 (0, 32) | 29 (0, 54) |
| 30 | 2 (0, 35) | 38 (0, 62) |
| 35 | 8 (0, 39) | 48 (0, 72) |
| 40 | 16 (0, 44) | 59 (19, 85) |
| 45 | 24 (0, 50) | 71 (41, 102) |
| 50 | 32 (0, 57) | 84 (59, 125) |
| 55 | 42 (0, 67) | 99 (73, 154) |
| 60 | 52 (0, 82) | 116 (85, 191) |
| 65 | 63 (24, 104) | 135 (97, 233) |
| 67 | 69 (37, 116) | 143 (102, 253) |
| 70 | 78 (51, 139) | 156 (109, 285) |
| 75 | 96 (65, 192) | 182 (124, 346) |
| 80 | 117 (76, 266) | 214 (141, 423) |
| 85 | 144 (88, 365) | 254 (163, 522) |
| 90 | 182 (105, 507) | 311 (194, 662) |
| 95 | 247 (133, 757) | 408 (247, 900) |
| 98 | 311 (161, 1002) | 506 (300, 1140) |
| 99 | 397 (197, 1331) | 635 (369, 1458) |

Posterior medians (and 95% credible intervals) are reported. When the required antibody level is predicted to be negative, 0 BAU/mL is reported, as negative antibody levels are not possible.

**Supplementary Table 8: Comparison of methods and results with previous studies considering waning vaccine efficacy against the Delta, Gamma and Omicron variants**

| Paper | Primary course vaccine | Booster vaccine | Dominant variant | Two-dose /booster /prior infection | Study design | Randomised vaccine /control assignment | Baseline comparator | Outcome | Location of result in paper | Group | Weeks since dose | Vaccine efficacy (%) |
|---|---|---|---|---|---|---|---|---|---|---|---|---|
| Katikireddi et al. (2022) [11] | ChAdOx1† | | Delta<br>Gamma | Two dose | Cohort*<br>Test-negative | x | Unvaccinated | Severe disease*<br>Symptomatic | Table 3 | Scotland (Delta) | 4-5<br>10-11<br>18-19 | 67.3 (65.3, 69.1)<br>59.3 (57.2, 61.4)<br>44.6 (41.5, 47.6) |
| | | | | | | | | | | Brazil (Gamma) | 4-5<br>10-11<br>18-19 | 68.4 (67.8, 68.9)<br>63.2 (62.2, 64.2)<br>57.7 (55.4, 60.0) |
| Menni et al. (2022) [12] | ChAdOx1†<br>BNT162b2*<br>mRNA-1273* | BNT162b2*<br>mRNA-1273* | Delta | Two dose<br>Booster dose* | Cohort | x | Unvaccinated | Any infection (mostly symptomatic) | (Figure 1A) Supp. Table 1 | All ages | 4<br>9<br>13<br>17<br>22<br>26 | 83.1 (82.2, 84)<br>79.3 (78.6, 80.0)<br>76.7 (76.0, 77.5)<br>75.8 (75.0, 76.4)<br>75.7 (74.9, 76.4)<br>75.2 (74.3, 76.1) |
| Horne et al. (2022) [13] | ChAdOx1†<br>BNT162b2* | | Delta | Two dose | Cohort | x | Unvaccinated | Any infection | Supp. Table 7 | Age 40-64 | 3-6<br>11-14<br>23-26 | 22 (19, 25)<br>−25 (−30, −20)<br>−86 (−79, −93) |
| | | | | | | | | | | Age 65+ | 3-6<br>11-14<br>23-26 | 57 (42, 68)<br>30 (20, 38)<br>−23 (−36, −11) |
| Andrews et al. (2022b) [10] | ChAdOx1†<br>BNT162b2* | BNT162b2<br>mRNA-1273<br>ChAdOx1†* | Omicron<br>Delta* | Two dose<br>Booster dose | Test-negative | x | Unvaccinated | Symptomatic | Table 3 | Two dose | 2-4<br>5-9<br>15-19<br>25+ | 48.9 (39.2, 57.1)<br>33.7 (25.0, 41.5)<br>17.8 (13.4, 21.9)<br>−2.7 (−4.2, −1.2) |
| | | | | | | | | | | BNT162b2 booster | 2-4<br>5-9<br>≥10 | 62.4 (61.8, 63.0)<br>52.9 (52.1, 53.7)<br>39.6 (38.0, 41.1) |
| | | | | | | | | | | mRNA-1273 booster | 2-4<br>5-9 | 70.1 (69.5, 70.7)<br>60.9 (59.7, 62.1) |
| Hogan et al. (2023) [14]‡ | ChAdOx1†<br>BNT162b2*<br>mRNA-1273* | BNT162b2*<br>mRNA-1273*<br>ChAdOx1†* | Omicron<br>Delta* | Two dose<br>Booster dose* | Modelling | x | Unvaccinated | Mild disease | Table 2 | All recipients | 13<br>26 | 12 (11.5, 12.4)<br>6.1 (5.8, 6.4)§ |
| Our Omicron analysis¶ | ChAdOx1† | | Omicron BA.4/5 | Two dose | Modelling | x | Vaccine non-responders | Any infection | Supp. Fig. 11 | All recipients (Scenario: Mean efficacy) | 5<br>10<br>20<br>26<br>29.9 | 18.5 (17.8, 19.2)<br>13.8 (13.3, 14.5)<br>7.6 (7.1, 8.2)<br>5.3 (4.8, 5.8)<br>4.1 (3.7, 4.7) |

We include studies of two doses of the ChAdOx1 nCoV-19 vaccine, and a study of efficacy after a booster dose, against the Delta, Gamma, and Omicron COVID-19 variants. Results are given with 95% confidence intervals or 95% credible intervals, depending on the paper. Results are presented for different age groups, where appropriate. *We do not report these results here, see the original paper. †ChAdOx1 refers to the ChAdOx1 nCoV-19 vaccine in this table. ‡Hogan et al. (2023) [14] fit a decay model to the results from Andrews et al. (2022b) [10]. The model does not appear to fit the data well - see Fig. 2 (top panel, mild disease) from their paper. §These results at 26 weeks were extrapolated from the model for timepoints beyond the data. ¶Our analysis predicts efficacy against Omicron by applying the relationship between anti-spike IgG levels and relative efficacy reported by Wei et al. [9] to the antibody trajectories predicted by our model.

### 5 Supplementary Figures

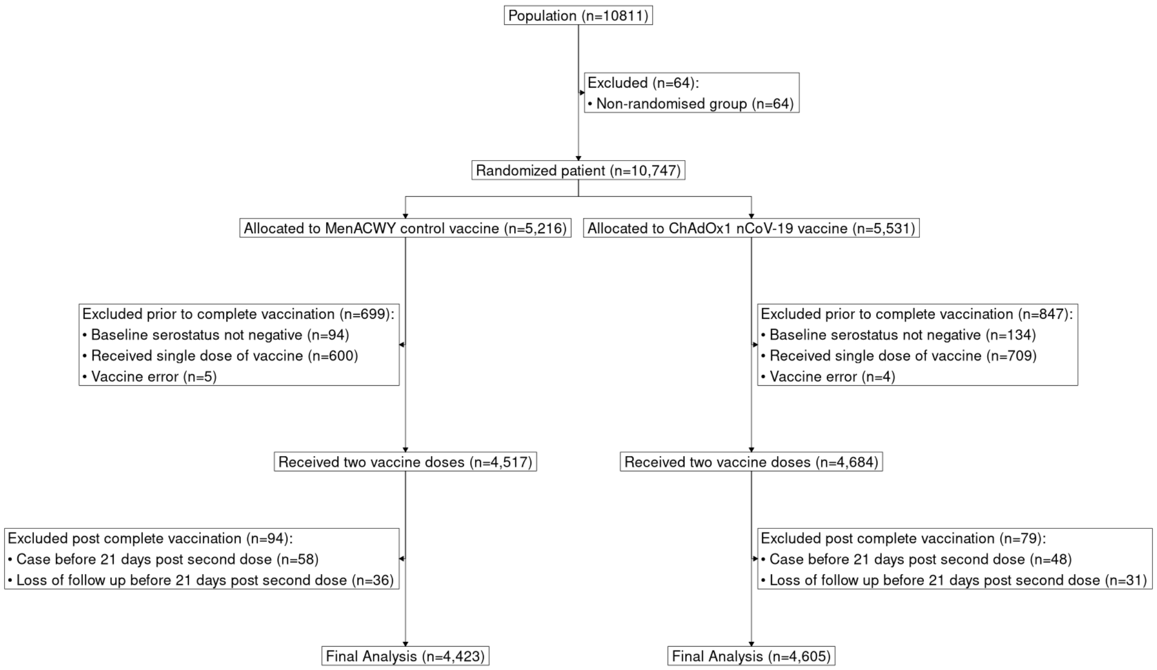

**Supplementary Figure 1: Flow chart showing inclusion of COV002 participants in analysis cohort.**

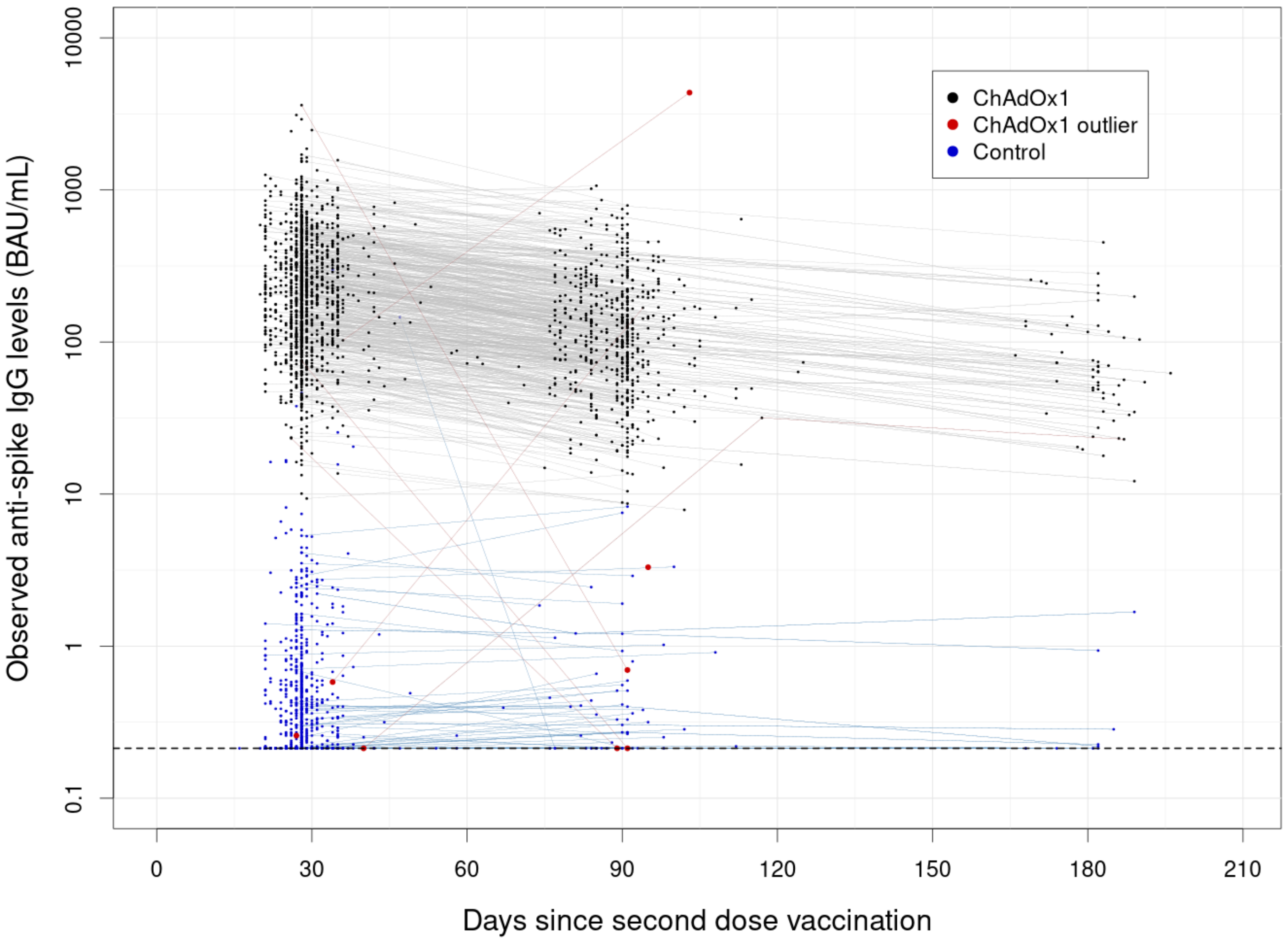

**Supplementary Figure 2: Observed anti-spike IgG antibody levels over time since vaccination, including outliers and measurements from control participants.** Each point represents an observation, and consecutive observations from the same individual are connected by a line. Black points denote included observations from vaccine arm participants. Red points denote observations excluded as outliers from vaccine arm participants. Blue points denote observations from control arm participants, which were not included in our model. When one of the points which a line connects is an outlier, a red line is used. Blue lines connect observations from control participants. A dotted horizontal line is drawn at the lower limit of quantification (LLOQ) 33 AU/mL (0.21 BAU/mL). Observations below the LLOQ are plotted at the LLOQ.

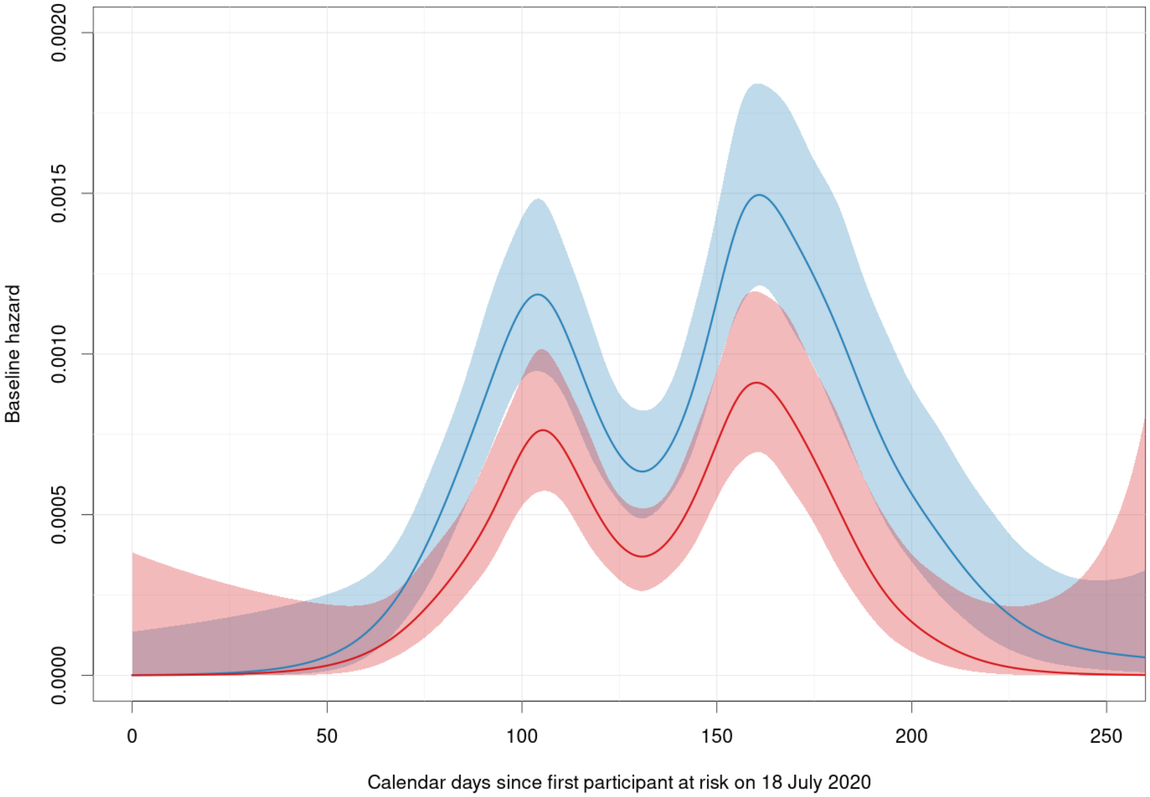

**Supplementary Figure 3: The estimated baseline hazard of infection in the trial by calendar days.** The (lower) red line and shaded region give the estimate and 95% confidence interval for baseline hazard of primary symptomatic COVID-19 infection. The (upper) blue line and shaded region give the estimate and 95% confidence interval for baseline hazard of any COVID-19 infection. The shaded regions overlap. Both were estimated from the control group only, using penalised restricted cubic splines in the R package survPen [15]. The baseline hazard is the hazard rate for an unvaccinated individual with . Few individuals had yet entered the study prior to 50 days since the start, and few remained in the study beyond 250 days since the start, hence the wide confidence intervals in those regions.

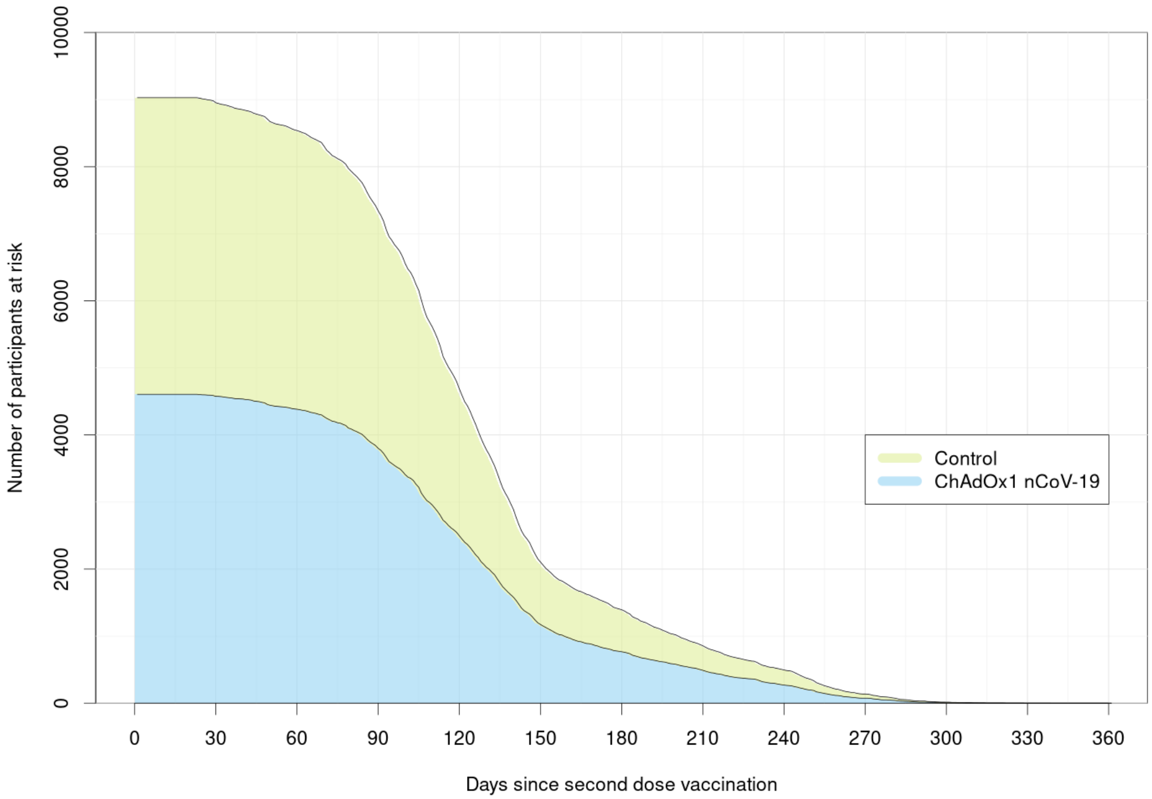

**Supplementary Figure 4: The number of individuals at risk over time since second dose in the vaccine and control arms.** The upper green area denotes the number of ChAdOx1 nCoV-19 (AZD1222) vaccinated participants at risk over time since second dose. The lower blue area denotes the number of control participants at risk over time since second dose. As such, the upper black line denotes the total number of participants at risk over time since second dose, and the lower black line the number of control participants at risk over time. Participants are censored from the analysis upon leaving the study, returning a positive COVID-19 test, and receiving a subsequent vaccination, among other reasons.

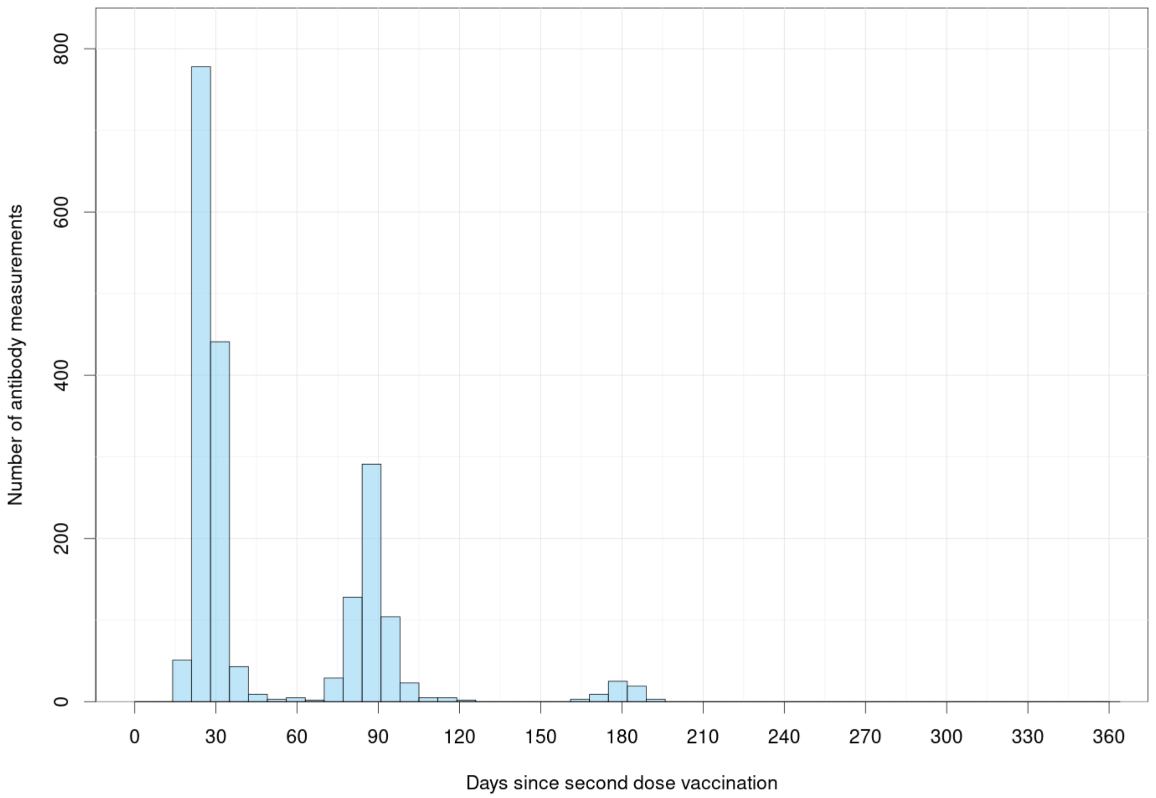

**Supplementary Figure 5: Histogram of the timings of anti-spike IgG samples since second dose vaccination.** Only anti-spike IgG samples from ChAdOx1 nCoV-19 (AZD1222) vaccinated participants are included. Samples from individuals more than 7 days prior to entering the at-risk period or less than 7 days before leaving the at-risk period are excluded. Hence, samples less than 7 days before returning a positive COVID-19 test, or after returning a positive test, are excluded.

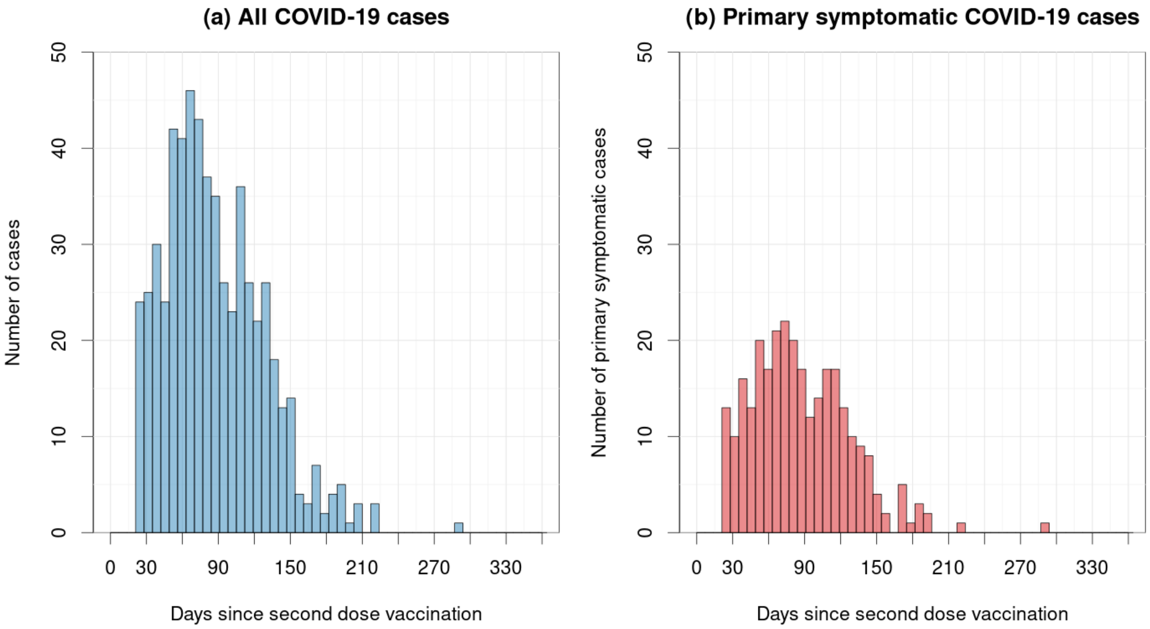

**Supplementary Figure 6: Histogram of the onset time since second dose of (a) any COVID-19 infection, (b) primary symptomatic COVID-19 infection.** (a) The blue histogram denotes the timings of the onset of any COVID-19 infection. (b) The red histogram denotes the timings of the onset of primary symptomatic COVID-19 infection.

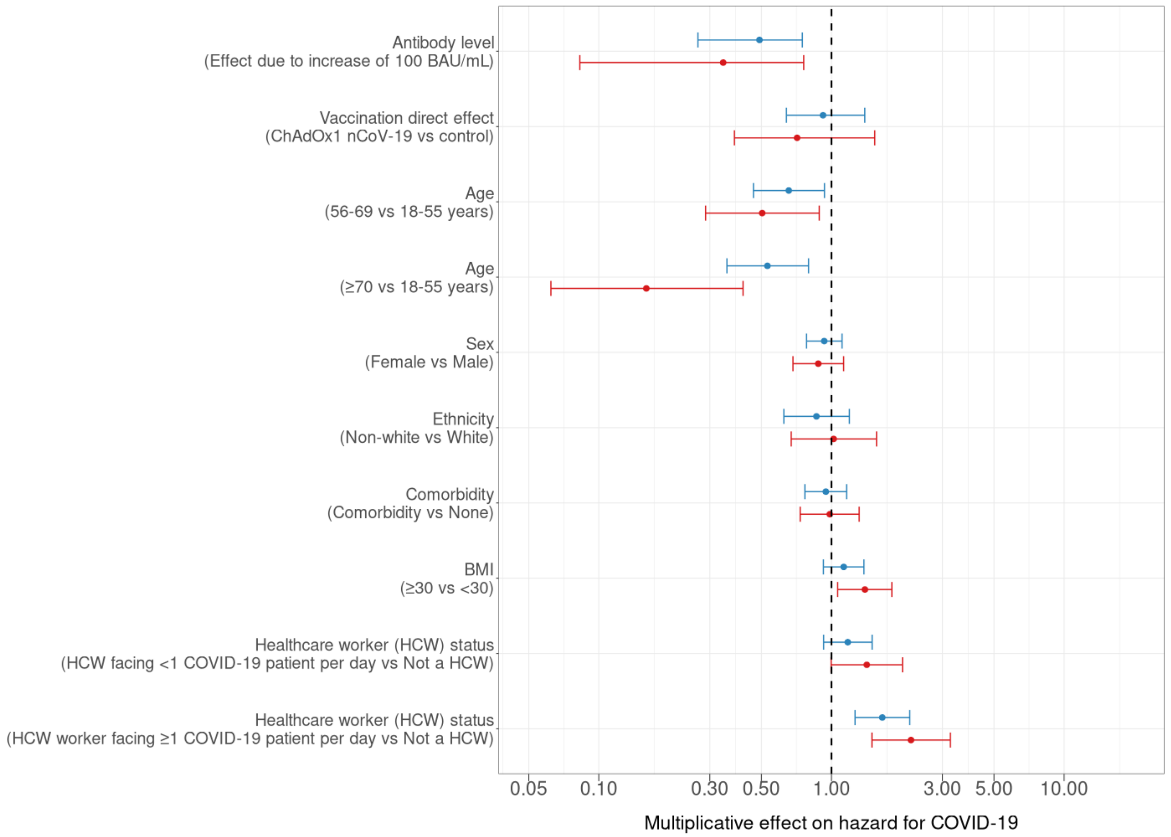

**Supplementary Figure 7: Covariate effects on hazard of COVID-19 infection.** The multiplicative effects due to different covariates on hazard of symptomatic COVID-19 infection and any COVID-19 infection. Points denote posterior medians and lines denote 95% credible intervals. Red denotes hazard against symptomatic infection and blue denotes hazard against any COVID-19 infection. These results are also given in Supplementary Table 6

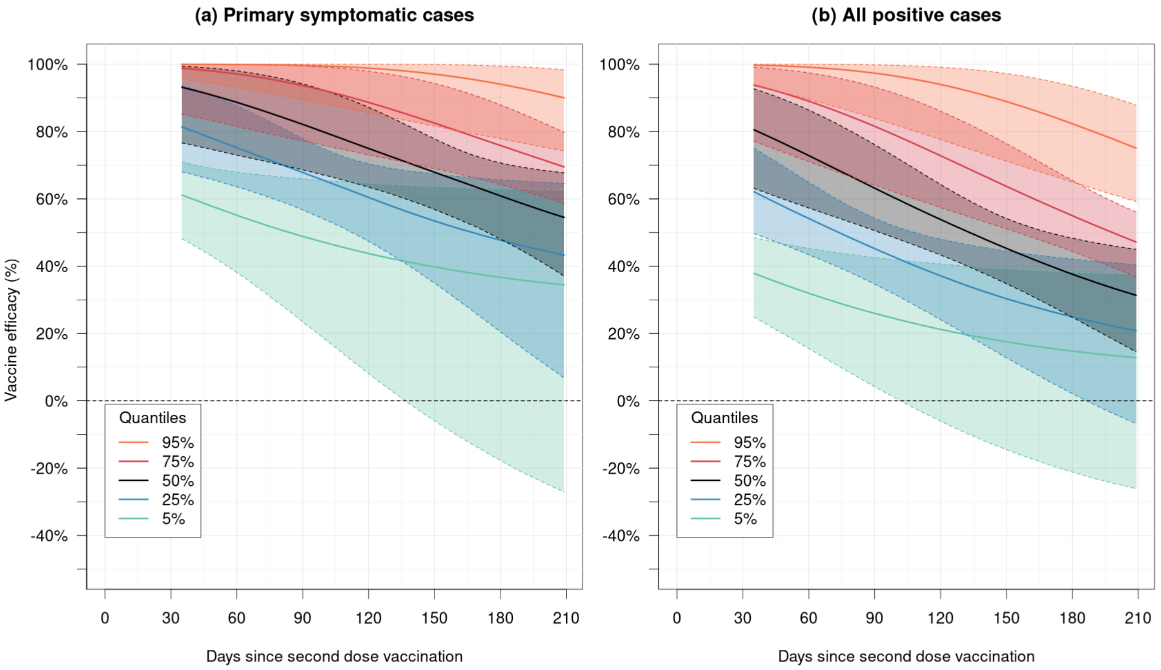

**Supplementary Figure 8: Quantiles of vaccine efficacy as a function of time since vaccination.** The curves show posterior medians and shaded regions 95% credible intervals. The 95%, 75%, 50%, 25% and 5% quantiles are plotted. (a) shows VE against symptomatic COVID-19 and (b) against all COVID-19 infection.

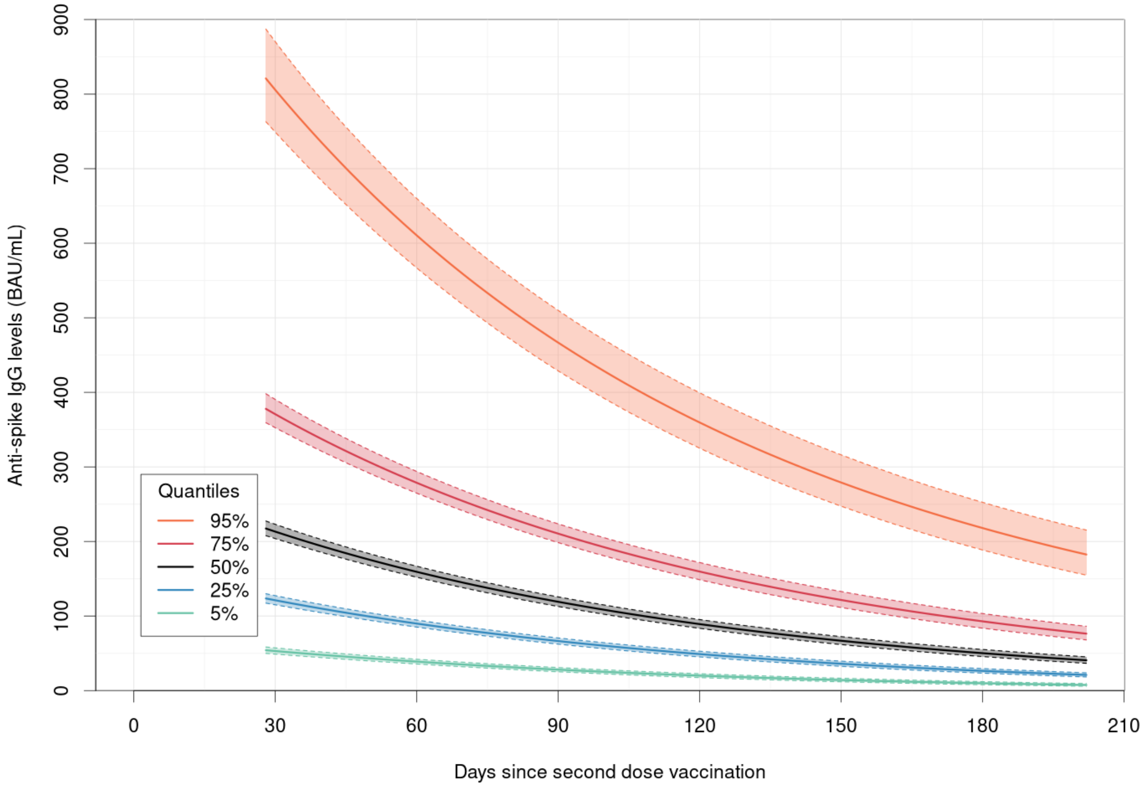

**Supplementary Figure 9: Quantiles of anti-spike IgG antibody levels as a function of time since vaccination.** The curves show posterior medians and shaded regions 95% credible intervals. The 95%, 75%, 50%, 25% and 5% quantiles are plotted.

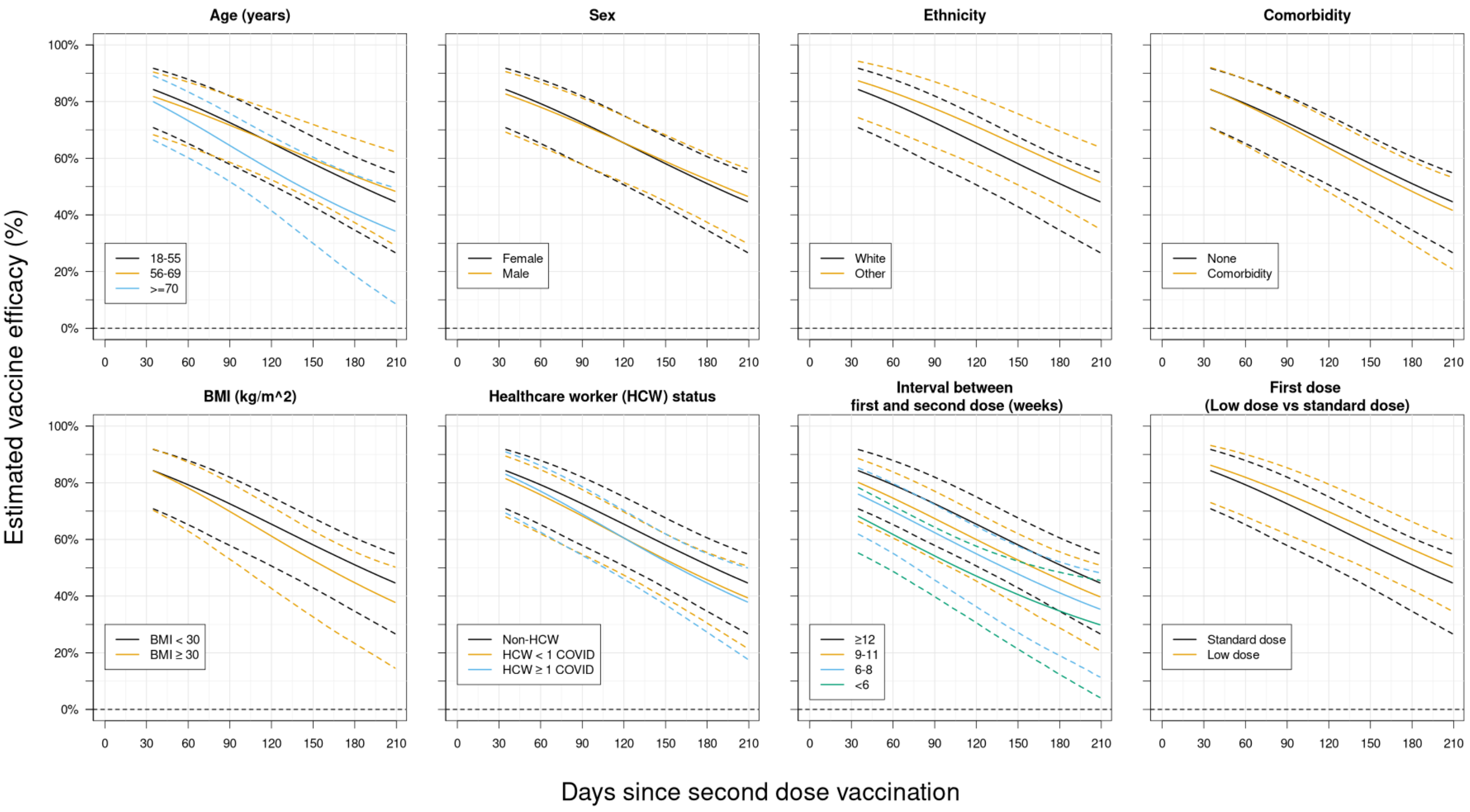

**Supplementary Figure 10: Vaccine efficacy against any COVID-19 infection plotted against time since vaccination for different covariate combinations.** The black lines represent an individual with "baseline" covariates: age 18-55, female, white ethnicity, no comorbidities, BMI<30 kg/m$^2$, >12 week interval between first and second dose, non-HCW, and receiving a standard dose as their first dose. All other lines represent an individual with one of the above covariates changed from these baseline values. The solid lines show posterior medians and dashed lines 95% credible intervals. "HCW < 1 COVID" refers to a healthcare worker facing <1 COVID-19 patient per day, "HCW $\geq$ 1 COVID" refers to a healthcare worker facing $\geq$1 COVID-19 patient per day.

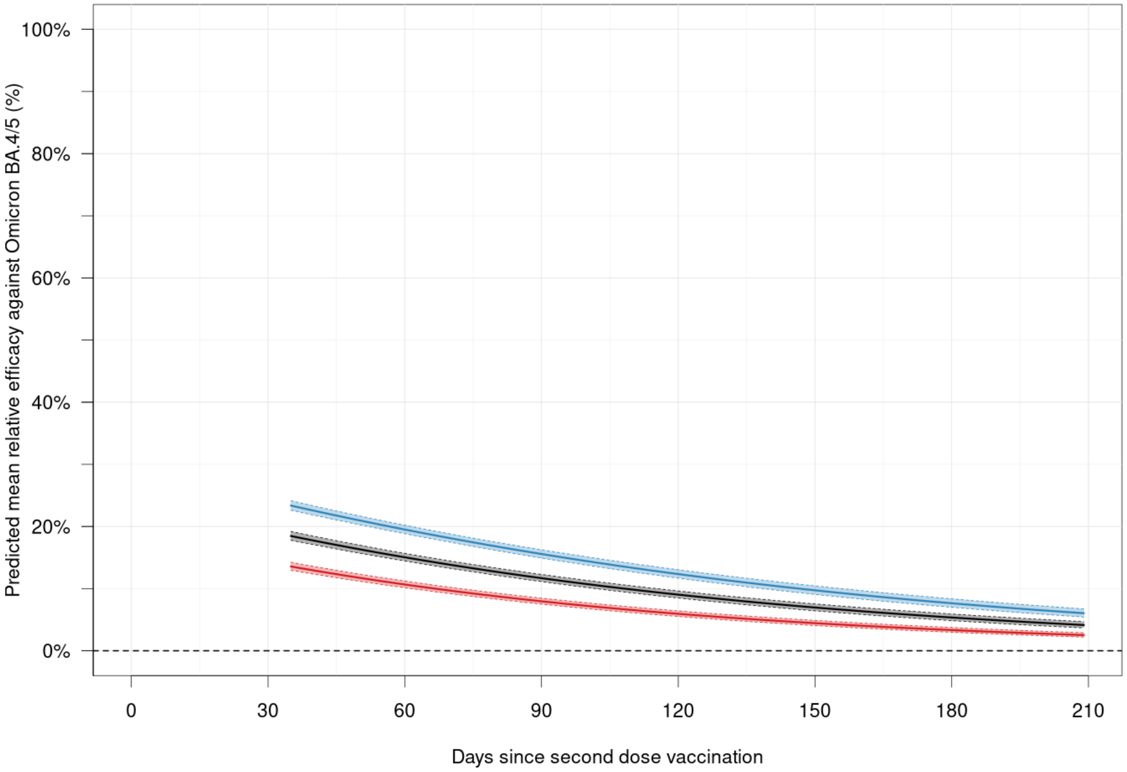

**Supplementary Figure 11: Predicted relative efficacy against SARS-CoV-2 Omicron BA.4/5 variant infection as a function of time since vaccination.** The black, blue and red lines represent estimates calculated using the reported posterior mean, and upper and lower 95% credible bounds for the relationship between relative efficacy and anti-spike IgG antibody levels from Wei et al. (2023). The solid lines show posterior medians and dashed lines 95% credible intervals, for the mean relative efficacy in the study.

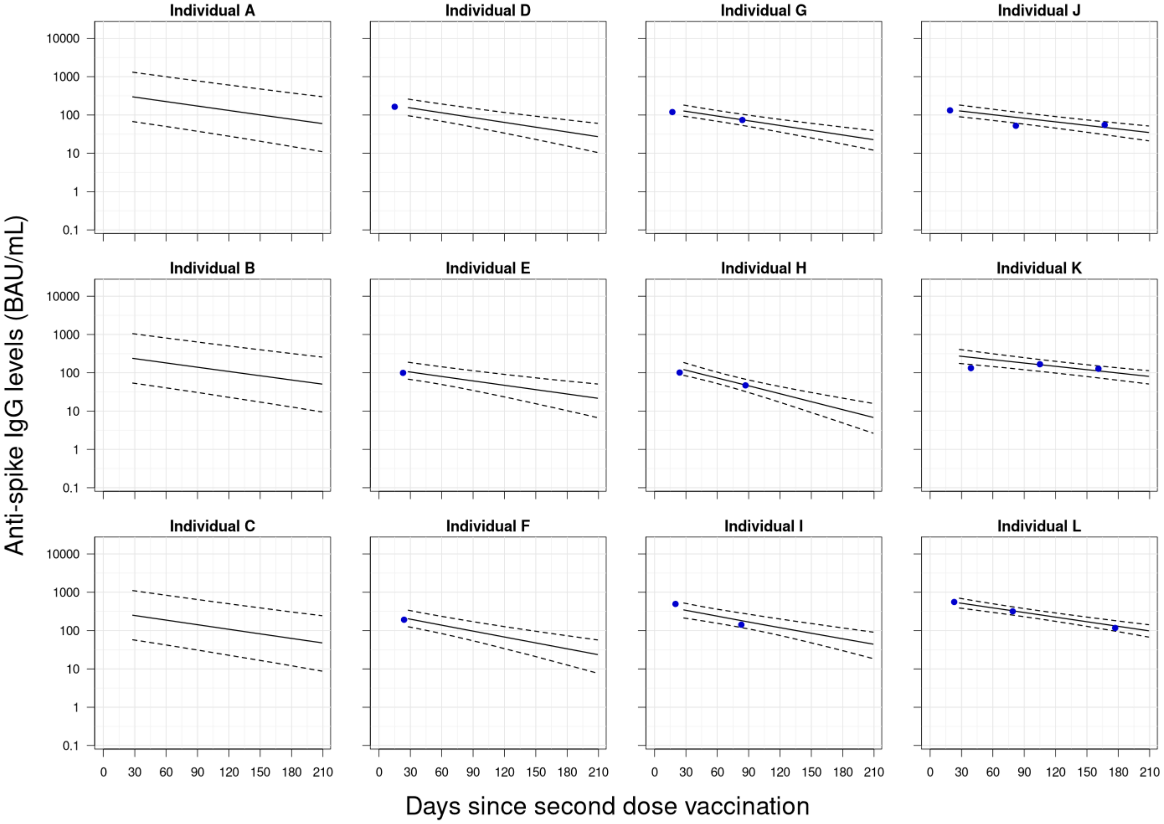

**Supplementary Figure 12: Estimated true anti-spike IgG levels for 12 randomly chosen individuals.** We randomly chose 3 individuals with 0, 1, 2 and 3 antibody observations available respectively, and plotted their estimated true anti-spike IgG levels over time. The solid line represents the posterior median, and the dashed lines represent a 95% credible interval. The blue points show the observed antibody levels. To be clear, the 3 plots on each row are 3 separate individuals (not the same individual with data consecutively being added).
